# Identification of rare loss of function variation regulating body fat distribution

**DOI:** 10.1101/2021.09.11.21263427

**Authors:** Mine Koprulu, Yajie Zhao, Eleanor Wheeler, Liang Dong, Nuno Rocha, Satish Patel, Marcel Van de Streek, Craig A. Glastonbury, Isobel D. Stewart, Felix R. Day, Jian’an Luan, Nicholas Bowker, Laura B. L. Wittemans, Nicola D. Kerrison, Debora M. E. Lucarelli, Inês Barroso, Mark I. McCarthy, Robert A. Scott, Vladimir Saudek, Kerrin S. Small, Nicholas J. Wareham, Robert K. Semple, John R. B. Perry, Stephen O’Rahilly, Luca A. Lotta, Claudia Langenberg, David B. Savage

**Author notes:** Correspondence to: Claudia Langenberg, David B. Savage. authors contributed equally (Mine Koprulu, Yajie Zhao). authors contributed equally (Claudia Langenberg, David B. Savage).

## Abstract

Biological and translational insights from large-scale, array-based genetic studies of fat distribution, a key determinant of metabolic health, have been limited by the difficulty in linking identified predominantly non-coding variants to specific gene targets. Rare coding variant analyses provide greater confidence that a specific gene is involved, but do not necessarily indicate whether gain or loss of function would be of most therapeutic benefit. Here we use a dual approach that combines the power of genome-wide analysis of array-based rare, non-synonymous variants in 184,246 individuals of UK Biobank with exome-sequence-based rare loss of function gene burden testing. The data indicates that loss-of-function (LoF) of four genes (*PLIN1, INSR, ACVR1C* and *PDE3B*) is associated with a beneficial impact on WHR_adjBMI_ and increased gluteofemoral fat mass, whereas *PLIN4* LoF adversely affects these parameters. This study robustly implicates these genes in the regulation of fat distribution, providing new and in some cases somewhat counter-intuitive insight into the potential consequences of targeting these molecules therapeutically.

## INTRODUCTION

Fat distribution is a heritable trait, commonly estimated by the relative amounts of waist and hip fat (waist-to-hip ratio, WHR) for a given body size. Genetic mechanisms linked to either relatively *lower* gluteofemoral or *higher* abdominal fat or both, have been shown to contribute to a greater WHR and its consistently adverse cardiometabolic consequences (1). Genome-wide array-based association studies have robustly identified many loci linked to WHR but thus far provided relatively limited biological and translational insights due to poor coverage of rare protein-coding variants and uncertainties connecting associated non-coding variants to functional genes (2, 3). Consequently, very few genes have been definitively linked to WHR and it is generally unknown whether a gain or loss of gene function is likely to drive observed associations.

The low frequency of rare (minor allele frequency [MAF] <0.5%, as defined by the 1000 Genomes Project (4)) functional variants which may have sizable effects on the encoded protein, may be a consequence of selective pressure acting against them, and previous studies have shown inverse relationships between allele frequency and effect size for complex traits (5, 6). Rare variants that occurred recently are also likely to be in low linkage disequilibrium (LD) with nearby common variants, facilitating fine-mapping and identification of causal variants and genes (7). However, rare variants are difficult to impute (8) so their study requires large, homogeneous samples and direct genotyping. To date, the vast majority of studies have explored the contribution of common variants in relation to WHR including the largest meta-analysis of imputed genome-wide association studies which included up to 694,649 individuals but only identified two variants at MAF 0.1-0.5% (3). The only other study which investigated the role of rare variants for WHR was a subsequent trans-ethnic Exomechip effort that identified 9 low frequency or rare variants with a lowest MAF of 0.1% (9).

The contribution of the full spectrum of rare variants to WHR using sequence data has not been studied, yet has the potential to provide a more direct link between gene and phenotype, and to facilitate translation from gene identification to drug development. Whilst the identification of coding variants in a specific gene clearly increases confidence in linking that particular gene to a trait, the impact of individual coding variants can still be very, or at least relatively, subtle. Individual variant testing, even using exome sequencing data in large populations, therefore still provides limited power and leaves residual uncertainty about the benefits of gain or loss of function of a particular gene. Exome wide scans of gene-based burden of rare loss-of-function variants have the potential to address this limitation (10-14). In this study, we use a dual approach that combines the power of large-scale genome-wide analysis of array-based rare, non-synonymous variants with exome-sequence-based rare gene burden testing to identify the putative function of variants, genes and pathways regulating body shape and fat distribution (assessed by BMI-adjusted WHR [WHR_adjBMI_]) and to determine their effects on body composition and metabolic health.

## RESULTS

A genome-wide analysis of directly genotyped, rare (0.1%≤MAF≤0.5%) non-synonymous variants associated with WHR_adjBMI_ at *p*<5×10^−8^ in 450,562 European ancestry individuals from UK Biobank identified lead variants in *PLIN1* p.L90P (rs139271800, EAF=0.1%), *PDE3B* p.R783X (rs150090666, EAF=0.1%), *ACVR1C* p.I195T (rs56188432, EAF=0.2%), *CALCRL* p.L87P (rs61739909; EAF=0.3%), *ABHD15* p.G147D (rs141385558; EAF=0.2%) and *PYGM* p.R50X (rs116987552, EAF=0.4%) (Figure 1, Supplementary Table 1). We observed a correlation of 0.99 and minor allele concordance of 0.99 comparing genotyped to whole-exome sequenced rare (0.1≤MAF≤0.5%) non-synonymous variants when testing the validity of rare, genotyped variants using exome-sequencing data from the overlapping samples (Supplementary Table 2).

**Figure 1.**
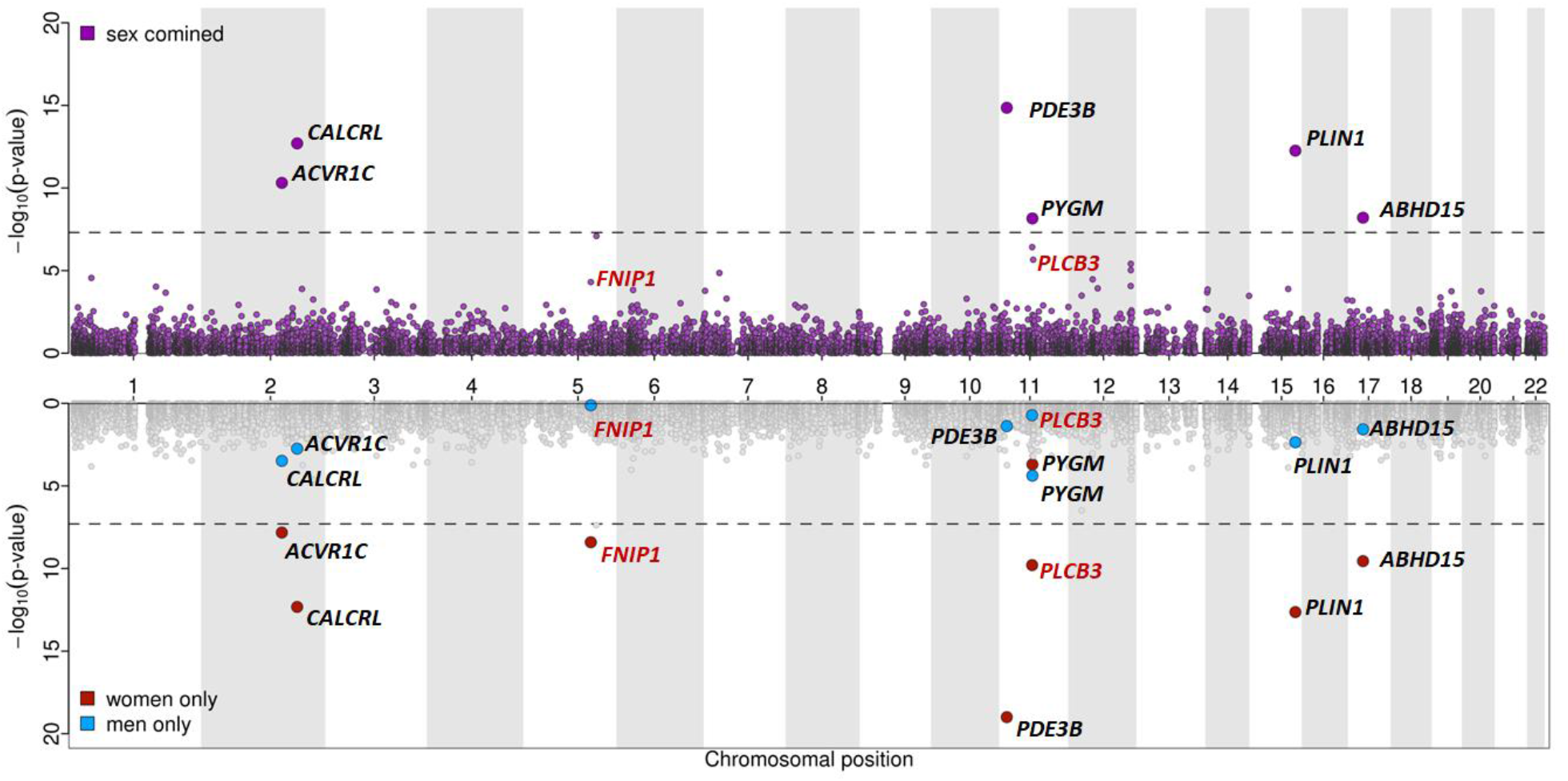
Miami plot for sex-combined and sex-specific single marker association results for WHR_adjBMI_. *Top*: Manhattan plot representing results from the main, sex-combined GWAS for WHR_adjBMI_ for genotyped, rare non-synonymous variants (0.1%≤MAF≤0.5%, correlation and rare allele concordance >0.9 when compared to the exome sequencing data). Gene annotations for the genome-wide significant variants from the main, sex-combined analyses are shown in black; gene annotations and significance from the main, sex-combined analyses for variants that were genome-wide significant in sex-specific analyses (women) only are shown in red. *Bottom*: Sex-specific significance of the variants highlighted above.

### Sex-differences in genetic effects of rare variants on WHR_adjBMI_

Common variant analyses have provided evidence of differences in genetic associations with fat distribution between men and women (3). In line with this, we found evidence of significant sex interactions, with stronger genetic effects in women, compared to men for all lead variants, except for *ACVR1C* p.I195T and *PYGM* p.R50X (Table 1). We therefore conducted sex-specific analyses which revealed two additional variants, *PLCB3* p.V806I (rs145502455, EAF=0.4%) and *FNIP1* p.R518Q (rs115209326, EAF=0.3%) to be genome-significant in (*p*<5×10^−8^) in women, with no effect in men (Table 1, Supplementary Table 1). No variants reached genome-wide significance in men only.

**Table 1:** Sex-stratified results for variants identified in joint and sex specific analyses of genotyped rare variants in UK Biobank. Unshaded rows present results discovered from the sex-combined analysis and grey shaded rows represent results from variants identified only in the sex-stratified (women) analysis. Abbreviations: *p* value Women, BOLT LMM p-value in women; *p* value Men, BOLT LMM p-value in men; Beta Women, effect size in women; Beta Men, effect size in men; SE Women, standard error in women; SE Men, standard error in men.

| Variant | rsID | Sex<br>interaction<br>*p* value | *p* value<br>Women | *p* value<br>Men | Beta<br>Women | Beta<br>Men | SE<br>Women | SE<br>Men |
| --- | --- | --- | --- | --- | --- | --- | --- | --- |
| ***PLIN1* p.L90P** | rs139271800 | $4.07 \times 10^{-3}$ | $1.90 \times 10^{-13}$ | $4.50 \times 10^{-3}$ | -0.273 | -0.126 | 0.039 | 0.042 |
| ***PDE3B* p.R783X** | rs150090666 | $5.66 \times 10^{-5}$ | $1.00 \times 10^{-19}$ | $4.10 \times 10^{-2}$ | -0.392 | -0.102 | 0.042 | 0.048 |
| ***ACVR1C* p.I195T** | rs56188432 | $3.61 \times 10^{-1}$ | $1.10 \times 10^{-8}$ | $3.40 \times 10^{-4}$ | -0.16 | -0.109 | 0.028 | 0.032 |
| ***CALCRL* p.L87P** | rs61739909 | $4.30 \times 10^{-3}$ | $4.30 \times 10^{-13}$ | $2.50 \times 10^{-3}$ | -0.171 | -0.082 | 0.024 | 0.026 |
| ***ABHD15* p.G147D** | rs141385558 | $4.53 \times 10^{-3}$ | $3.40 \times 10^{-10}$ | $3.80 \times 10^{-2}$ | 0.168 | 0.057 | 0.026 | 0.029 |
| ***PYGM* p.R50X** | rs116987552 | $3.56 \times 10^{-1}$ | $2.90 \times 10^{-4}$ | $5.40 \times 10^{-5}$ | 0.079 | 0.095 | 0.021 | 0.024 |
| ***PLCB3* p.V806I** | rs145502455 | $2.34 \times 10^{-2}$ | $1.60 \times 10^{-10}$ | $2.00 \times 10^{-1}$ | 0.126 | 0.029 | 0.021 | 0.024 |
| ***FNIP1* p.R518Q** | rs115209326 | $6.09 \times 10^{-4}$ | $4.80 \times 10^{-9}$ | $8.40 \times 10^{-1}$ | -0.128 | -0.003 | 0.023 | 0.026 |

### Genomic context and fine-mapping analyses

We found strong statistical evidence for causal associations of rare non-synonymous variants in *PLIN1, PDE3B, ACVR1C* and *CALCRL* (Supplementary Note 1, Supplementary Table 3, Supplementary Figure 1) through conditional analysis and fine-mapping, whereas genomic context analyses did not support the causality of the identified rare lead variants in *ABHD15* or *PYGM* from the joint (sex-combined) analysis, and of *PLCB3* and *FNIP1* in the women-only analysis (Supplementary Note 2, Supplementary Table 4). Bioinformatic analysis of these variants strongly predicted that the *PDE3B* variant p.R783X would truncate *PDE3B* within the catalytic site, impairing *PDE3B* catalytic activity if expressed (Supplementary Note 3), whereas predictions of the functional impact of the *PLIN1, ACVR1C* and *CALCRL* variants were less conclusive (Supplementary Note 3).

### Exome-sequenced based burden testing of rare, loss-of function variants

Next, we considered the genes identified in the single variant analysis for exome-sequence-based gene rare LoF and missense burden testing in 184,246 individuals in UK Biobank (see Methods, Gene-based association testing) and found that *PLIN1, PDE3B, ACVR1C*, and *CALCRL* were all significantly associated with lower WHR_adjBMI_ at a Bonferroni corrected threshold (*p*<0.0125). Predicted loss of function (pLoF) variants showed the most significant association for *PLIN1* and *PDE3B*, moderate impact variants for *CALCRL*, and the combination of pLoF with moderate impact variants for *ACVR1C* (Supplementary Table 5).

In order to identify additional genes where loss of function may regulate fat distribution, we extended this approach to a hypothesis free, exome-wide analysis (*p*<2.53×10^−6^) for WHR_adjBMI_ using more stringent quality control (QC) parameters (see Methods). This identified *PLIN4* and *INSR* in at least one of the variant categories (see Methods), in addition to *PLIN1, ACVR1C* and *PDE3B* (Figure 2, Supplementary Table 6). *PLIN4, INSR* and *PDE3B* all showed significantly larger standardized effect sizes for women compared to men (*p*<0.05) in gene-based analyses, in line with the single marker results (Supplementary Table 7). While the joint effect of rare LoF variants in *PLIN4* (65 variants, 1,065 carriers) was associated with a higher WHR_adjBMI_ (Beta = 0.16 [0.10 – 0.22], *p*=5.86×10^−7^), the combination of rare LoF variants in *PLIN1* (31 variants, 393 carriers) was associated with a lower WHR_adjBMI,_ (Beta = -0.27 [-0.17 – -0.36], *p*=9.82×10^−9^) (Supplementary Table 6). The lead *PLIN1* LoF variant (*PLIN1* p.T338DfsX51, rs750619494) is predicted to result in a frameshift from amino acid 338 with a premature stop at amino acid 388, though it may well be subject to nonsense mediated RNA decay. Several additional *PLIN1* variants are similarly expected to result in early truncations or nonsense mediated RNA decay (Supplementary Table 8). In either instance, these variants are expected to impair Plin1 interaction with *ABHD5* and thus it’s regulation of adipose triglyceride lipase (*ATGL*) (15). In the case of *PLIN4* (p.Q372X, rs201581703), the variant list also included early frameshift/premature stop variants predicted to result in nonsense mediated RNA decay.

**Figure 2.**
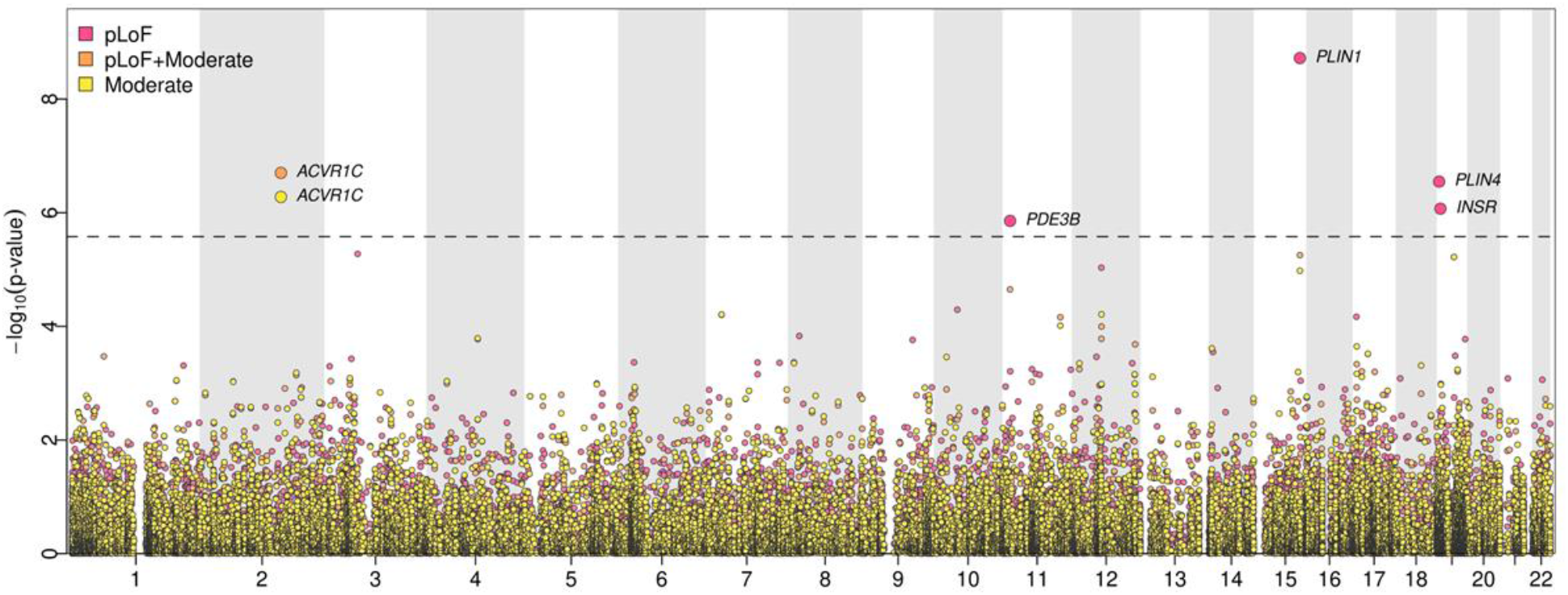
Gene-based association results. Gene-based exome-wide discovery results for WHR_adjBMI_. The horizontal dashed line represents the exome-wide significance threshold (*p*=2.53×10^−6^).

We next assessed phenotypic associations with refined measures of fat distribution and cardiometabolic parameters and diseases. Bioelectrical Impedance Analysis (BIA) derived body fat compartment measurements (16) showed that *PLIN4* (pLoF) was associated with higher android and trunk fat, and lower gynoid and leg fat (Figure 3, Supplementary Figure 2, Supplementary Table 9) whereas *PLIN1* (pLoF) acted in the opposite direction. Fat distribution is strongly linked to insulin resistance, but as direct indicators of insulin resistance are not currently available in UK Biobank, we evaluated the impact of these genes on metabolic indicators typically associated with insulin resistance (17, 18) (Figure 3, Supplementary Figure 2, Supplementary Table 9). *PLIN4* LoF was associated with higher triglycerides (TGs), TG/HDL (triglyceride/high-density lipoprotein cholesterol) ratio and higher HbA1c levels. The associations for *PLIN1* consistently contrasted with those of *PLIN4* with lower TGs, TG/HDL ratio and additionally higher HDL cholesterol levels, in keeping with a beneficial impact on insulin sensitivity. In keeping with these findings, *PLIN4* LoF was nominally associated with an increased risk for type 2 diabetes (T2D) (OR=1.36 [1.06-1.66], *p*=0.04) in the Type 2 Diabetes Knowledge Portal (T2DKP; https://t2d.hugeamp.org/), though none of the genes showed a significant association with T2D in UK Biobank through our analysis or in the AstraZeneca PheWAS Portal (https://azphewas.com/) (Supplementary Figure 2; Supplementary Table 9; Supplementary Table 10). *PLIN1* LoF showed nominal significance for a lower risk of cardiovascular heart disease (CHD) (*p*=0.03, OR=0.55[0.31-0.91]) in our analysis of UK Biobank, a finding supported by association between *PLIN1* LoF and reduced susceptibility to chronic ischemic heart disease (OR=0.40 [0.32-0.75], *p*=4.49×10^−4^) in the AstraZeneca PheWAS Portal (14).

**Figure 3.**
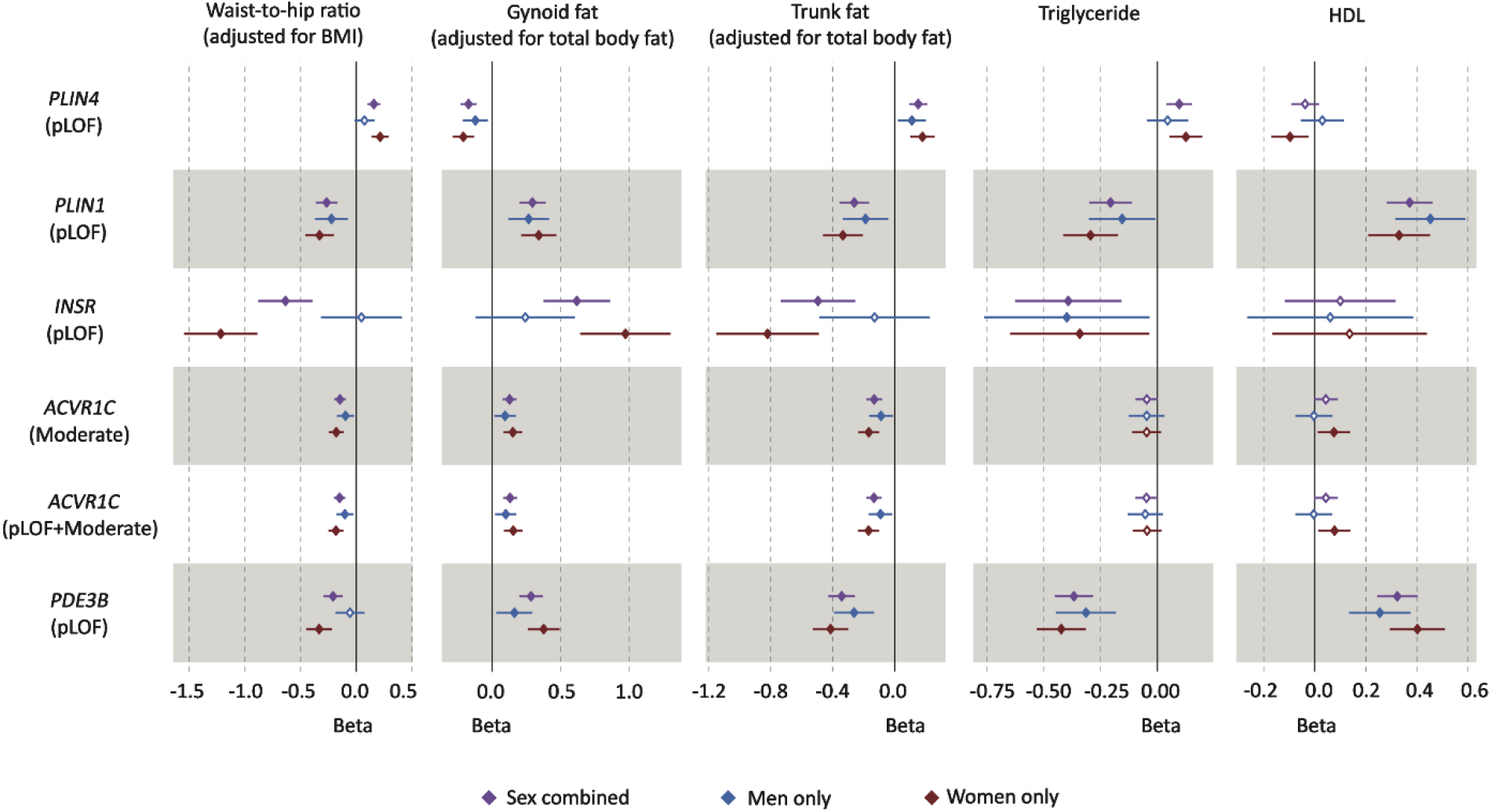
Forest plot of phenotypic associations for significant variant-gene categories. Black represents the sex-combined, blue represents the men-only and red represents the women-only analysis. Horizontal lines represent 95% confidence intervals. Waist-to-hip ratio adjusted for BMI (n=184,246), gynoid fat adjusted for total body fat (n= 178,143), trunk fat adjusted for total body fat (178,143), triglyceride levels (n=175,271) and HDL cholesterol (n=161,239) were all driven from UK Biobank (See Supplementary Table 12 for details).

Similarly to *PLIN1*, the combined effect of LoF variants in the *INSR* (27 variants, 61 carriers) was associated with lower WHR_adjBMI_ (Beta=-0.64[-0.39 – -0.88], *p*=6.21×10^−7^; Supplementary Table 6). Although a few common intronic variants (rs1035942, rs1035940, rs62124511, rs34194998) and a low-frequency synonymous variant (rs1799815) in the *INSR* have previously been associated with WHR_adjBMI_ (3, 19), the causal mechanism underlying these associations remains unknown. Our gene-based findings indicate that the *lNSR* can alter body fat distribution through loss of function. Given the fact that the *INSR* gene encodes the insulin receptor itself and that both biallelic and heterozygous loss of function variants in this gene have long been linked with monogenic severe insulin resistance syndromes (20), this evidence for loss of function of the *INSR* having a seemingly beneficial effect on fat distribution i.e. lower WHR_adjBMI_ is surprising. Importantly, none of the *INSR* mutations previously linked to monogenic disease were present in our UK Biobank analysis. The lead *INSR* variant (p.R525X) is predicted to result in truncation of the protein within the extracellular domain preventing interaction of the extra- and intra-cellular domains, and thus formation of a functional receptor. In the homozygous state, this would be expected to lead to monogenic severe insulin resistance. In terms of body fat distribution, heterozygous *INSR* LoF was also associated with higher gynoid and leg fat, and lower android and trunk fat mass (Figure 3, Supplementary Figure 2, Supplementary Table 9). Similarly to the cardio-metabolic associations of *PLIN1* indicating a beneficial effect, heterozygous loss of *INSR* was associated with lower TGs and a lower TG/HDL ratio (Figure 3, Supplementary Figure 2, Supplementary Table 9). It was also associated with lower LDL (low-density lipoprotein) cholesterol levels but was not associated with altered HDL (Figure 3, Supplementary Figure 2, Supplementary Table 9). Despite these seemingly favourable changes in fat distribution and plasma lipids, *INSR* LoF showed a nominal association for increased susceptibility to T2D in the T2DKP (OR=3.67[2.50-4.83], *p*=0.02) (Supplementary Table 10).

For *ACVR1C*, the genetic architecture of gene-based results was slightly different, gene-based association results were significant for (i) the combined burden of pLoF and moderate impact variants and (ii) for moderate impact variants only. There were 130 rare moderate impact variants and 9 rare high impact variants included in this analysis [1414 and 16 carriers, respectively]. The combined effect of pLoF and moderate impact variants and moderate impact only variants were both associated with lower WHR_adjBMI_ (Beta=-0.15 [-0.10 – -0.20] and -0.15[-0.10 – -0.20], *p*=1.68×10^−7^ and 4.57×10^−7^, respectively; Supplementary Table 6). In this instance, the highest-ranking variant was the previously reported p.I195T variant (19, 21). In silico predictions including M-CAP (22), REVEL (23), SIFT (24), PolyPhen-2 (25) and PROVEAN (26) all asses this variant to be damaging to the protein and structural modelling also suggests that it is likely to have a sizable impact (Supplementary Note 3). CADD (27) also estimates this variant to be among the top 1% of the deleterious variants ranked by CADD (score = 27.1). To test this prediction, we performed a luciferase reporter assay in HEK293 cells which strongly suggested that the mutation impairs *ACVR1C* signalling (Figure 4).

**Figure 4.**
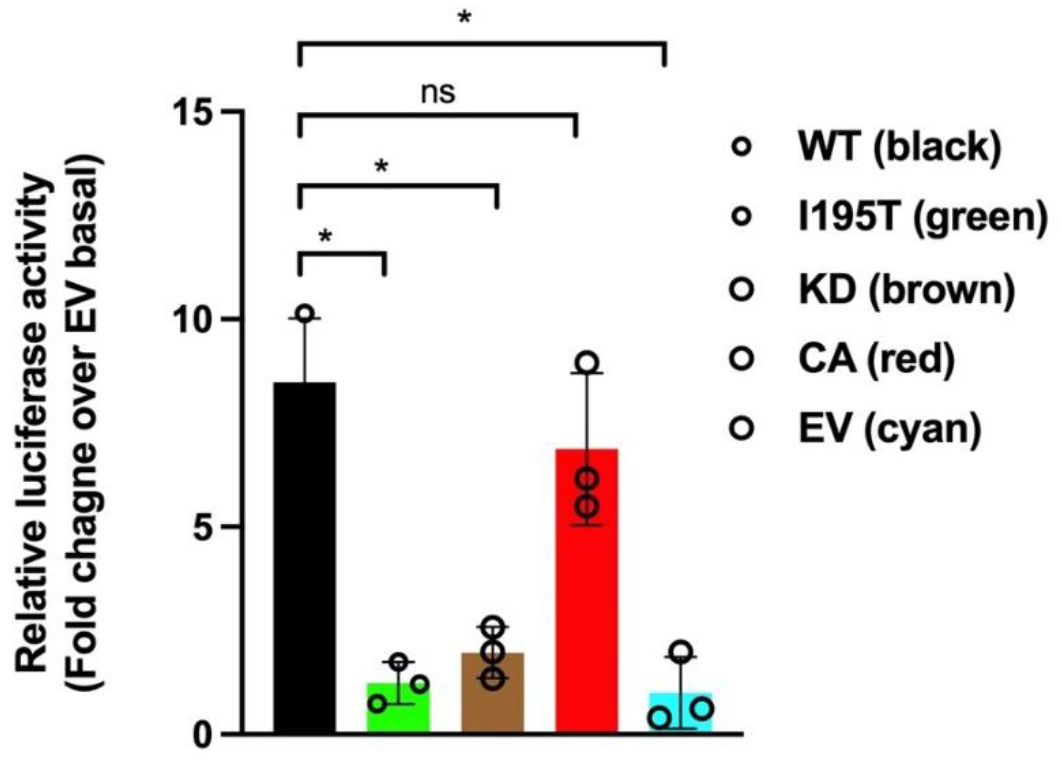
Functional impact of *ACVR1C* I195T variant on Smad signalling. HEK293 cells were transiently transfected with ACVR1C expression constructs and their receptor components, along with firefly and *Renilla* luciferase expression plasmids. Firefly luciferase activity was normalised to *Renilla* activity and the luciferase activity in non-stimulated cells transfected with empty vector (EV) was set to 1. A constitutively active (CA) *ACVR1C* variant T194D and a kinase dead (KD) variant K222R were included for comparison. Results from three independent experiments are presented as mean ± SD. Statistical significance was evaluated by one-way ANOVA with Tukey’s post hoc test for multiple comparisons between pairs. WT, wild type ACVR1C. * **P** < 0.001, ns not significant.

The phenotypic associations for *ACVR1C* LoF for fat categories were similar to *PLIN1* LoF, with significant associations with higher gynoid and leg fat, and lower android and trunk fat. However, the cardio-metabolic associations were less clear for *ACVR1C* with an association with lower TG and but not HDL or the TG/HDL ratio (Figure 3, Supplementary Figure 2; Supplementary Table 9).

Finally, the combined effect of LoF variants in *PDE3B* was also significantly associated with lower WHR_adjBMI_. However, leave-one-out analysis suggested that this association was mainly driven by the premature stop variant (p.R783X, rs150090666; *p*-value after dropping the variant=0.49; Supplementary Table 11). All other candidate genes remained at least nominally significant after dropping the most significant variant (Supplementary Table 11). Our analysis of the *PDE3B* p.R783X variant was in line with previous reports associating it with lower triglyceride levels and higher HDL (28, 29). *PDE3B* p.R783X has also been reported to be associated with higher apolipoprotein B, lower apolipoprotein A1 levels and other haematological traits (30). This variant was reported to be significantly associated with cardiovascular disease when meta-analysed in UK Biobank and other cohorts (29, 31).

## DISCUSSION

Central adiposity has long been linked to insulin resistance and metabolic disease (32-37) but exactly why this is the case and, other than sex hormones, what exactly determines fat distribution remains incompletely understood. So, what have we learnt from human genetics thus far? Firstly, that inheritance contributes to WHR (3, 38). Secondly, monogenic partial lipodystrophies indicate that single gene variants can be sufficient to mediate substantial changes in fat distribution, classic examples being mutations in *LMNA* and *PPARG*. Interestingly, both proteins are expressed in all white adipocytes and yet specific loss of function variants are consistently associated with loss of hip and leg fat whereas visceral fat is preserved (39). Thirdly, whilst the beneficial impact of thiazolidinediones, one of very few drugs that clearly improve insulin sensitivity, was recognised before the discovery of *PPARG* mutations in patients with partial lipodystrophy, this link attests to the potential for human genetics to inform drug discovery. Fourthly, GWAS studies have identified many loci associated with WHR (2, 3, 9) though these have yet to be translated into therapeutic targets. Finally, Mendelian Randomization has been used to establish that genetic mechanisms linked to greater WHR_adjBMI_ can be causally linked to the risk of cardiovascular disease and type 2 diabetes through either relatively *lower* gluteofemoral or *higher* abdominal fat or both (40). Furthermore, these associations are very likely to be underpinned by insulin resistance as the genetic risk score for WHR_adjBMI_ was also shown to be strongly associated with elevated fasting insulin, higher triglycerides and lower HDL cholesterol (40).

Our WHR_adjBMI_ single variant analysis in samples from 450,562 UK Biobank participants revealed missense variants in 3 genes (*CALCRL, PLIN1* and *ACVR1C*) and a nonsense variant in *PDE3B*. All these genes are highly expressed in adipose tissue in keeping with emerging evidence that adiposity itself is largely centrally mediated whereas where excess energy is stored is regulated within adipose tissue itself (2, 41). *CALCRL* is also expressed in a host of other tissues and its role in adipose tissue remains to be established (42, 43). *PLIN1* is a lipid droplet surface protein almost exclusively expressed in adipocytes and has a well-established role in regulating both triglyceride and diacylglycerol hydrolysis (44). *PDE3B* is expressed in many tissues but has long been linked to adipocyte lipolysis, and specifically to insulin mediated inhibition of lipolysis (45, 46). Several lines of evidence have recently implicated *ACVR1C* in the regulation of lipolysis, but it is expressed in many tissues in addition to adipose tissue and further work is required to convincingly establish exactly what it does in adipocytes (47-49).

In terms of the impact of the specific mutations present in each gene, the *PDE3B* p.R783X is clearly expected to impair *PDE3B* catalytic activity, and thus potentially to increase cAMP levels and lipolysis, but the impact of the other three variants is far less certain (see Supplementary Note 3). Interestingly, gene based LoF burden analyses for all four genes were at least nominally significant, suggesting that the single variants were most likely to impair function of the encoded proteins. At least when transiently transfected into cultured cells, our functional data is also consistent with the LoF predictions for the *ACVR1C* p.I195T variant.

Our exome wide analyses confirmed significant effects of LoF variants in *PLIN1, ACVR1C* and *PDE3B*. In the cases of *ACVR1C* and *PDE3B*, the lead LoF variants were the same as single variants reported above, namely the *ACVR1C* p.I195T and *PDE3B* p.R783X variants. The fact that the phenotypic associations of the *PLIN1* p.L90P variant are directionally consistent with the *PLIN1* LoF gene burden data suggests that this variant is likely to be a loss of function variant too. Both the lead *PLIN1* variant and several additional *PLIN1* variants are expected to result in early truncations or nonsense mediated RNA decay. These data are consistent with Laver *et al*.’s assertion that *PLIN1* haploinsufficiency is not associated with lipodystrophy (50). Interestingly, several heterozygous *PLIN1* frameshift variants had previously been linked to partial lipodystrophy (51). None of these variants overlap with those identified in UK Biobank to date and none are predicted to result in nonsense mediated RNA decay. Instead, in several cases, immunoblotting of adipose tissue lysates confirmed expression of an elongated form of Perilipin 1 in addition to the wildtype copy, so perhaps expression of these mutant forms with an altered carboxy-terminus, accounts for the seemingly ‘opposite’ phenotypes (51, 52).

Gene burden testing also highlighted the role of *PLIN4* LoF variants in fat distribution, though in this case, these had an adverse impact on WHR_adjBMI_. Whilst a higher WHR_adjBMI_ would conventionally be deemed to be metabolically adverse, it is possible that this need not be the case for all genetic perturbations, however, our phenotypic analyses were consistent with the predicted outcomes for all the above genes, in fact, the phenotypic associations for *PLIN1* and *PLIN4* were consistently opposite. Similar to *PLIN1, PLIN4* is highly expressed in adipose tissue, but it is also expressed in heart and skeletal muscle, and the *PLIN4* knockout mouse has not been reported to have an adipose tissue phenotype to date (53).

The last gene identified in the exome wide gene burden analysis was the *INSR*. In this instance, the lead variant (p.R525X) is expected to truncate the protein in the alpha subunit shortly before the disulphide bond normally connecting alpha and beta subunits. This is expected to abrogate synthesis of functional receptor. Even if truncated protein were synthesised this would not be able to dimerise with or exert dominant negative activity over co-expressed wild type receptor, and so heterozygosity for the truncating variant would be expected to reduce functional receptor protein by ∼50%. In keeping with this, biallelic mutations in this domain usually cause extreme IR classified as Donohue or Rabson-Mendenhall syndrome. Parents of affected children have not been systematically studied and are generally held to be metabolically normal. In contrast, heterozygous *INSR* variants in the intracellular beta subunit, which are synthesised and interfere with wild type receptor function, cause type A insulin resistance (20). Whilst fat mass is often reduced in Donohue’s syndrome, heterozygous variants associated with type A insulin resistance are not reported to be associated with fat redistribution and interestingly do not typically lead to fatty liver or dyslipidaemia (20). Our data suggest, surprisingly, that the *INSR* LoF variants favourably impact WHR_adjBMI_ and LDL cholesterol. Whilst the *INSR* LoF association with T2D was relatively weak statistically and not seen in all the cohorts assessed, it is conceivable that *INSR* LoF might adversely affect pancreatic beta cell function and/or insulin sensitivity despite the apparently beneficial impact on fat distribution. The change in triglycerides is somewhat reminiscent of the well described absence of dyslipidaemia in patients with monogenic severe insulin resistance due to bi- or mono-allelic *INSR* mutations, so again this does not preclude the *INSR* LOF variants being associated with reduced insulin sensitivity.

Our sex-specific analyses consistently revealed stronger effects in women than in men. These data are consistent with the fact that WHR is more strongly associated with insulin resistance in women than in men (54). Fat mass in women is consistently significantly higher than men of a similar BMI, who typically have higher lean/muscle mass. The adverse impact of a lack of lower limb/gluteofemoral fat on metabolism is strikingly apparent in patients with familial partial lipodystrophy, particularly types 2 and 3, due to specific mutations in *LMNA* and *PPARG* respectively (39). In these and in fact in all forms of partial lipodystrophy, metabolic disease manifests considerably earlier and is typically more severe in women than in men (39, 55, 56).

Our analyses have several limitations which future work should help to resolve. Firstly, the statistical power to detect associations, particularly when examining rare variants, depends on the sample size. Hence, there is the opportunity to discover additional findings when the WES data is released in the full UK Biobank cohort or other large-scale studies. Secondly, the phenotypic follow up of cardiometabolic diseases for the candidate genes was primarily conducted in UK Biobank, a population cohort with a limited number of cases of specific diseases. Our follow-up in T2DKP revealed the potential of datasets enriched for cases to increase statistical power in phenotypic follow ups. Thirdly, fat distribution is strongly associated with insulin resistance, but the UK Biobank cohort did not provide fasting samples so direct measures of insulin and inferred indices of insulin sensitivity are not available.

In conclusion, our analysis strongly implicates at least four genes in the regulation of fat distribution. Furthermore, the data suggests that inhibitors of *PLIN1, PDE3B* and *ACVR1C* might favourably impact fat distribution and associated metabolic phenotypes whereas *PLIN4* inhibition is likely to have adverse health consequences. The data in *PLIN1* needs to be tempered by the earlier reports linking some specific *PLIN1* LoF variants with partial lipodystrophy. Finally, the data on the *INSR* seemed to suggest a potential disconnect between an apparently favourable impact of LoF variants on WHR and an apparently adverse impact on T2D risk. These findings provide valuable insight into the potential of these genes as therapeutic targets.

## METHODS

### The UK Biobank Resource

The UK Biobank is a large-scale prospective population-based study of approximately 500,000 participants aged 40 – 69 at the time of recruitment (57). Recruitment took place between 2006 – 2010 in centres across the United Kingdom and participants have deep phenotypic information collected from initial and repeat assessment visits, health records, self-reported survey information, linkage to death and cancer registries, urine and blood biomarkers and other phenotypic endpoints. A Seca 200 cm tape was used to measure waist and hip circumference at the baseline visit, and BMI was calculated from height and weight measurements. WHR_adjBMI_ was constructed as the ratio of waist and hip circumferences adjusted for age, age^2^ and BMI (measured at the baseline assessment visit). Residuals were calculated for men and women separately and then transformed using the rank-based inverse normal transformation. All additional phenotypes are described in Supplementary Table 12.

### Genome-wide association scan of genotyped rare nonsynonymous genetic variants

Genetic variants were genotyped in UK Biobank using the Affymetrix UK BiLEVE or the Affymetrix UK Biobank Axiom arrays (57). Genotyping underwent quality control procedures including (a) routine quality checks carried out during the process of sample retrieval, DNA extraction, and genotype calling; (b) checks and filters for genotype batch effects, plate effects, departures from Hardy Weinberg equilibrium, sex effects, array effects, and discordance across control replicates; and (c) individual and genetic variant call rate filters as previously described (57). We further excluded genetic variants with a genotype call rate below 95% and variants that were (i) not rare (0.1%≤MAF≤0.5%) or (ii) not non-synonymous or (iii) had poor correlation (r<0.9) or rare allele concordance (<0.9) when compared to the whole exome sequence data (Supplementary Table 2). Genomic annotations were performed using the ANNOVAR software (58). The coordinates of genotyped rare variants were lifted over from GRCh37 to GRCh38 using liftOver and all reported positions in this study are in GRCh38. A total of 13,181 genetic variants in 7,481 genes were available for analysis. Genome-wide association analyses were performed using the BOLT-LMM software (59) in 450,562 participants of European Ancestry defined using a K-means clustering approach applied to the first four principal components calculated from genome-wide SNP genotypes.

Sex-specific genome-wide association analyses were performed using the BOLT-LMM software (59) in 206,082 men and 244,478 women of European ancestry from UK Biobank. Evidence for sex differences at the variants identified in the sex combined analysis were formally tested in unrelated individuals using a linear regression model including an interaction term between the genetic variant and sex using the same covariates used in the discovery analysis.

### Conditional analyses and fine-mapping

At each associated genomic region, we conducted systematic analyses of the genomic context of associations. Our goal was to establish whether or not the identified rare nonsynonymous variants are likely to be the causal variants for the association with WHR_adjBMI_. At each region 1 Mb either side of the nonsynonymous genetic variants associated with WHR_adjBMI_, we conducted both approximate and formal conditional analyses. We considered the association of all genetic variants in the regions regardless of functional annotation or allele frequency using directly-genotyped and imputed data (imputed using the Haplotype Reference Consortium (HRC) and UK10K haplotype resource). First, approximate conditional analyses were conducted on summary-level estimates using GCTA (60, 61) to identify sets of conditionally-independent index genetic variants (p<5×10^−8^). Individual-level genotypes for the conditionally-independent variants identified in this first step were then extracted in 350,721 unrelated European ancestry participants of UK Biobank and their independent association was confirmed in multivariable linear regression models including all variants put forward from approximate analyses. Then, at each region, we statistically decomposed the identified index signals by conditioning on the other conditionally-independent index variants. We then performed Bayesian fine-mapping (62) to estimate the posterior probability of association for each variant (PPA, where 0% indicates that the variant is not causal and 100% indicates the highest possible posterior probability that the variant is causal) and define the 99% credible set at that signal (i.e. a set of variants in a genomic window that accounts for 99% of the PPA at that association signal). To generate credible sets, the association results at each locus were converted to Bayes factors (BF) for each variant within the locus boundary. The posterior probability that a variant-j was causal was defined by:

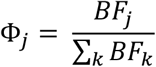

where, BF_j_ denotes the BF for the j_th_ variant, and the denominator is the sum of BF_s_ for all included variants at that signal. A 99% credible set of variants was created by ranking the posterior probabilities from highest to lowest and summing them until the cumulative posterior probability exceeded 0.99 (i.e. 99%).

### UK Biobank exome-sequence data processing and QC

Whole exome sequencing (WES) data of 200,643 UK Biobank participants made available in October 2020 were downloaded in VCF and PLINK formats. The details of the UK Biobank WES data processing are provided in detail elsewhere (63, 64). Further data processing and quality control has been described previously (65). In brief, we did not apply additional QC based on QUAL (variant site-level quality score, Phred scale) or AQ measures (variant site-level allele quality score reflecting evidence for each alternate allele, Phred scale). Site-level filtering was applied for targeted biallelic calls if the AB ratio (no. of alternate allele reads/total depth) was ≤0.25 or ≥0.8. Variant-level QC filters were applied if any of the variants had (i) genotype missingness > 5%, (ii) maximum read depth (DP) of less than 10 across samples or (iii) had GQ less than 20 for over 20% of the calls. After applying these filters, 7.3% of the variants were flagged as poor quality and not taken forward for further analysis.

### Variant annotation and definition of gene burden sets

We annotated variants released in UK Biobank 200K whole exome sequencing VCF files in build hg38, using the Variant Effect Predictor (VEP) tool release 99 provided by Ensembl (66). In addition to default VEP features such as the consequence and impact of the variant, overlapping gene, position at cDNA and protein level and codon and amino acid change, if applicable, we have used the following plugins for annotation: (i) SIFT (67), which predicts whether an amino acid substitution affects protein function based on sequence homology and the physical properties of amino acid, (ii) Polyphen-2 (25), which predicts possible impact of an amino acid substitution on the structure and function of a human protein, (iii) CADD (27) which provides deleteriousness prediction scores for all variants based on diverse genomic features and (iv) LOFTEE (68)which provides loss of function prediction for variants. We annotated each variant using the most severe consequence across overlapping transcripts in Ensembl. We defined loss of function variants as those with ‘high’ impact predict by VEP. This includes frameshift variants, transcript ablating or transcript amplifying variants, splice acceptor or splice donor variants, stop lost, start gained or stop gained variants. ‘Moderate impact’ variants include missense variants, in-frame deletion or insertions, missense variants and protein altering variants.

### Gene-based association testing

In our discovery stage, we used the method STAAR (variant-Set Test for Association using Annotation infoRmation), which is a computationally scalable method for very large whole-exome sequence (WES) and whole-exome sequence (WGS) studies and large-scale biobanks. STAAR uses a Generalized Linear Mixed Model (GLMM) framework that includes linear and logistic mixed models and can also account for both relatedness and population structure for both quantitative and dichotomous traits (69). In our analysis, we used the genotype dosage matrix as the genotype input and covariates including age at first check (age), age^2^, sex, genotyping array, top ten genetically derived principal components (PC1-PC10) generated from the SNP array data, exome sequencing batch and the sparse GRM. For rank-based inverse normal transformed WHR_adjBMI_, we also added BMI as a covariate. We excluded the samples from our analysis if they did not pass UK Biobank quality control parameters, were non-European ancestry or if they withdrew consent from the study (n=184,246) (65).

We ran STAAR with its default options without additional functional annotations. For each gene, with at least two variants with MAF ≤ 0.5%, we conducted gene-based association analysis for the following three variant categories: rare variants predicted by VEP to be a) loss of function (pLoF; i.e. high impact), b) missense (Moderate; i.e. moderate impact) or c) both (pLoF+Moderate). For each variant clustering of a gene, STAAR will provide p-values for several collapsing burden tests including SKAT (sequence kernel association test), Burden test, and ACAT-V (set-based aggregated Cauchy association test). In addition, the output of STAAR also includes the omnibus p-value (STAAR-O) by using the combined Cauchy association test to aggregate the association across the different tests.

After identifying the genes with STAAR-O p-value over the threshold for exome-wide significance (*p*<2.53×10^−6^), we applied more stringent QC filters on the genotype calls of the included variants. We set to missing genotype calls which did not meet the following QC criteria: 1. Genotype Quality (GQ) ≥ 20 for heterozygous variants; 2. Depth (DP) ≥ 7 for SNVs and DP≥ 10 for InDels; 3. A binomial test on allelic balance using the Allelic Depth (AD) FORMAT field for heterozygous variants with *p*≥1×10^−3^. We then repeated the STAAR analysis using the filtered genotype dosage matrix.

To examine the extent to which the gene-based association is driven by single variants, we conducted a sensitivity leave-one-out analysis for each significant gene (*p*<2.53×10^−6^), testing the significance of the gene-based association after excluding each variant.

### Secondary association testing

We created dichotomous dummy variables using the filtered genotype dosage matrix for each identified gene, where samples with one or more rare alleles were set as “1” and the samples without rare alleles were set as “0” for different variant clustering settings of each gene. Then we combined these dummy variables into a single file and transformed it to BGEN format, which was used as the genotype input for association testing using a linear mixed model implemented in BOLT-LMM to account for cryptic population structure and relatedness (59). The GRM in BOLT-LMM was generated from the autosomal genetic variants that were common (MAF > 1%), passed quality control in all 106 batches, and were present on both genotyping arrays (65). Covariates included age, sex, and PC1-PC10, genotyping chip and exome sequencing batch. For rank-based inverse normal transformed WHR_adjBMI_, covariates also included BMI. We excluded the same group of samples as we did for STAAR analyses.

To test for heterogeneity of effect sizes between men and women for significant genes identified in the gene-based analyses, we used a Z-test to compare effect size estimates for each gene calculated in the sex-specific analyses.

### Phenotypic associations

The gene-based phenotypic associations using the same STAAR and BOLT-LMM pipelines for the following continuous phenotypes: BMI, BIA-derived gynoid fat, BIA-derived leg fat, BIA-derived android fat, BIA-derived trunk fat, BIA-derived arm fat, triglyceride levels, HDL cholesterol, LDL cholesterol, HbA1c levels (see Supplementary Table 12 for phenotype details). Body fat compartments were predicted using bioimpedance measurements in UK Biobank. The details for the prediction of body fat compartments in UK Biobank are described elsewhere (16).

We have also investigated gene-based phenotypic associations for binary disease outcomes: type 2 diabetes and cardiovascular heart disease. As BOLT-LMM is based on the linear mixed model which cannot give an accurate effect estimate for binary variables, we have also applied a generalized linear model (GLM) to estimate the Odd Ratio (OR) for binary phenotypes. We also looked up these binary outcomes in other resources such as the AstraZeneca PheWAS Portal (https://azphewas.com/, accessed on 02/09/2021) (14) and the Type 2 Diabetes Knowledge Portal (T2DKP; https://t2d.hugeamp.org/, accessed on 02/09/2021). The AstraZeneca PheWAS Portal also uses UK Biobank as their primary resource, but have access to a larger dataset of 281,104 exomes. We looked up results for T2D (N cases = 1,671; N controls = 160,949) and chronic ischaemic heart disease (defined by ICD-10 code I25; N cases =24,147, N controls = 176,170). In the T2DKP, we also looked up results for T2D (N=43,125).

### ACVR1C dual-luciferase assay

HEK293 cells were seeded at a density of 150,000 cells per well in 24-well tissue culture plates pre-treated with poly-D-lysine. On the following day, medium was replaced with Opti-MEM I Reduced Serum medium and a total of 550 ng of plasmid DNA; this included different pcDNA3.1 based ACVR1C constructs listed in the table below, as well as constructs encoding receptor components (ACVR-IIB and CRIPTO) along with firefly (consisting of the SMAD binding elements) and *Renilla* (control) luciferase reporter plasmids. Lipofectamine 3000 Reagent was used for the transfection according to the manufacturer’s protocol. Opti-MEM I Reduced Serum medium was then replaced with DMEM growth medium 6 hours post transfection.

| pcDNA3.1 construct | Receptor components |
| --- | --- |
| WT | ACVR-IIB, CRIPTO, pGL4.48[*luc2P*/SBE/Hygro], pRL-SV40 |
| I195T | ACVR-IIB, CRIPTO, pGL4.48[*luc2P*/SBE/Hygro], pRL-SV40 |
| K222R | ACVR-IIB, CRIPTO, pGL4.48[*luc2P*/SBE/Hygro], pRL-SV40 |
| T194D | ACVR-IIB, CRIPTO, pGL4.48[*luc2P*/SBE/Hygro], pRL-SV40 |
| EV (empty vector) | ACVR-IIB, CRIPTO, pGL4.48[*luc2P*/SBE/Hygro], pRL-SV40 |

The dual-luciferase reporter assay was performed according to the manufacturer’s protocol (Promega, USA). Cells were washed once with DPBS, followed by an active lysis procedure. Briefly, 125 µl of passive lysis buffer was added in each well and the cells were subjected to one cycle of a freeze-thaw process. Cell lysates were cleared of cell debris by centrifugation at 21,130 *x g* for one minute. The assay was conducted in a 96-well plate format. In each assay, 20 µl of cleared supernatant was pre-dispensed, followed by sequential measurement of firefly and *Renilla* luciferase using a Tecan Spark 10M plate reader (Tecan, Switzerland). Firefly luciferase activity was normalised for *Renilla* luciferase activity, and then further normalised with values from non-stimulated cells transfected with empty pcDNA3.1 vector (EV). We also studied a constitutively active (ACVR1C p.T194D) mutant and a kinase dead (ACVR1C p.K222R) mutant for comparison (70, 71). The experiment was repeated with fresh transfections on three separate occasions.

## Data Availability

This research has been conducted using the UK Biobank resource (application no. 44448 and 9905). Access to the UK Biobank genotype and phenotype data is open to all approved health researchers (http://www.ukbiobank.ac.uk/).

http://www.ukbiobank.ac.uk/

## Acknowledgements

This research has been conducted using the UK Biobank resource. Access to the UK Biobank genotype and phenotype data is open to all approved health researchers (http://www.ukbiobank.ac.uk/). This study was funded by the United Kingdom’s Medical Research Council through grants MC_UU_12015/1, MC_PC_13046, MC_PC_13048 and MR/L00002/1. This work was supported by the MRC Metabolic Diseases Unit (MC_UU_12012/5) and the Cambridge NIHR Biomedical Research Centre and EU/EFPIA Innovative Medicines Initiative Joint Undertaking (EMIF grant: 115372). R.K.S, D.B.S. and S.O’R. are supported by the Wellcome Trust (WT 210752, WT 219417 and WT 214274 respectively) the MRC Metabolic Disease Unit, the National Institute for Health Research (NIHR) Cambridge Biomedical Research Centre and the NIHR Rare Disease Translational Research Collaboration. K.S.S. is supported by MRC Project Grant L01999X/1. Some computation was enabled through access granted to K.S.S. to the MRC eMedLab Medical Bioinformatics infrastructure, supported by the Medical Research Council (grant number MR/L016311/1). M.McC. is a Wellcome Senior Investigator supported by Wellcome grants 098381, 090532, 106130, 203141. M.McC. declares that the views expressed in this article are those of the authors and not necessarily those of the NHS, the NIHR, or the Department of Health; he has served on advisory panels for Pfizer, Novo Nordisk and Zoe Global, has received honoraria from Merck, Pfizer, Novo Nordisk and Eli Lilly, and research funding from Abbvie, Astra Zeneca, Boehringer Ingelheim, Eli Lilly, Janssen, Merck, Novo Nordisk, Pfizer, Roche, Sanofi Aventis, Servier, and Takeda. M.K. is supported by the Gates Cambridge Trust. The authors gratefully acknowledge the help of the MRC Epidemiology Unit Support Teams, including Field, Laboratory and Data Management Teams.

## Author contributions

Data collection and analysis: M.K., Y.Z., E.W., L.D., N.R., S.P., M.V.S., C.G., I.D.S, F.R.D, J.L., N.B., L.B.L.W., N.D.K., V.S.,

Study supervision: D.M.E.L., I.B., M.I.McM., R.A.S., K.S.S., N.J.W., R.K.S., L.A.L, J.R.B.P., S.O’R., C.L., D.B.S.

