## Supplementary Materials for "Identification of rare loss of function variation regulating body fat distribution"

### Table of Contents

Supplementary Table 1: Variants associated with waist-to-hip ratio adjusted for body mass index ($WHR_{adjBMI}$) in the discovery genome-wide analysis of rare non-synonymous variants. ... 1

Supplementary Table 2. a. Correlation between the UK Biobank WES data and the genotype data for rare nonsynonymous variants with a minor allele frequency (MAF) between 0.1% - 0.5% in overlapping samples. b. Box plot of rare allele concordance between the UK Biobank WES data and the genotype data for rare nonsynonymous variants with a MAF between 0.1% - 0.5% in overlapping samples. ... 2

Supplementary Table 3: a. Conditionally independent index variants and fine-mapping at the *CALCRL*, *PLIN1*, *PDE3B* and *ACVR1C* loci. b. Formal conditional analyses at the *CALCRL, PLIN1, PDE3B* and *ACVR1C* loci. ... 3

Supplementary Table 4: Conditionally-independent index variants at the *ABHD15, PYGM, PLCB3 and FNIP1* regions. ... 5

Supplementary Table 5: STAAR-O (2) gene-based results for genes discovered in the single variant analysis. ... 6

Supplementary Table 6: Significant gene-based results from the exome-wide scan for $WHR_{adjBMI}$. ... 7

Supplementary Table 7: Sex-specific results for gene-based analysis. ... 8

Supplementary Table 8: Quality control measurements for variants included in the refined gene-based tests and single marker association results for all included variants. ... 9

Supplementary Table 9: a. Summary statistics for quantitative traits from gene-based association analyses in sex-combined, women-only and male-only data. b. Summary statistics for binary traits from gene-based association analyses in sex-combined, women-only and male-only data. Details for all phenotypes and are given on Supplementary Table 12. ... 9

Supplementary Table 10: a. Lookup from Type 2 Diabetes Knowledge Portal for type 2 diabetes. ... 10

Supplementary Table 11: Leave one out analysis results for the most significant single variant in significant genes. ... 14

Supplementary Table 12. Phenotypes used for phenotypic associations in UK Biobank. ... 15

Supplementary Figure 1: Regional association plots of the overall and statistically-decomposed signals at the *CALCRL*, *PLIN1*, *PDE3B* and *ACVR1C* genes. ... 17

Supplementary Figure 2: Association of the significant genes with other body fat distribution or cardiometabolic trait related phenotypes in UK Biobank for (a) continuous phenotypes and (b) binary phenotypes. There was no rare predicted loss of function variant carriers for *INSR* in coronary heart disease (CHD) cases in women-only analyses. Abbreviations: pLoF, predicted loss of function; WHRadjBMI; waist-to-hip ratio adjusted for body mass index; BMI, body mass index; TG, triglycerides; HD; high density lipoprotein, LDL low-density lipoprotein; TG/HDL, triglyceride to high density lipoprotein ratio; T2D, type 2 diabetes; CHD, coronary heart disease. ... 18

**Supplementary Table 1: Variants associated with waist-to-hip ratio adjusted for body mass index ($WHR_{adjBMI}$) in the discovery genome-wide analysis of rare non-synonymous variants.**

| Gene | dbSNP rsID | Genomic coordinate, chromosome and position | Effect allele and other allele | Effect allele frequency in % (minor allele count) | Protein change (amino acid corresponding to the effect allele) | Beta (SE) for $WHR_{adjBMI}$ per effect allele in univariate analysis, in SD units | p-value | Beta (SE) for $WHR_{adjBMI}$ per effect allele in conditional analyses*, in SD units | p-value conditional analyses* |
|---|---|---|---|---|---|---|---|---|---|
| *Sex-combined* | | | | | | | | | |
| *PLIN1* | rs139271800 | chr15:89671546 | G and A | 0.1 (1,127) | p.L90P (P) | -0.21 (0.029) | $5.5\times10^{-13}$ | -0.21 (0.029) | $5.5\times10^{-13}$ |
| *PDE3B* | rs150090666 | chr11:14843853 | T and C | 0.1 (932) | p.R783X (X) | -0.26 (0.032) | $1.4\times10^{-15}$ | -0.25 (0.032) | $6.2\times10^{-15}$ |
| *ACVR1C* | rs56188432 | chr2:157550353 | G and A | 0.2 (2,113) | p.I195T (T) | -0.14 (0.021) | $4.9\times10^{-11}$ | -0.14 (0.021) | $5.4\times10^{-12}$ |
| *CALCRL* | rs61739909 | chr2:187380712 | G and A | 0.3 (2,954) | p.L87P (P) | -0.13 (0.018) | $2.0\times10^{-13}$ | -0.12 (0.018) | $5.9\times10^{-12}$ |
| *ABHD15* | rs141385558 | chr17:29566527 | T and C | 0.3 (2,441) | p.G147D (D) | 0.11 (0.019) | $6.3\times10^{-9}$ | 0.07 (0.019) | 0.00019 |
| *PYGM* | rs116987552 | chr11:64759751 | A and G | 0.4 (3,764) | p.R50X (X) | 0.09 (0.016) | $7.0\times10^{-9}$ | 0.06 (0.015) | 0.00037 |
| *Sex-specific analysis in women* ‡ | | | | | | | | | |
| *PLCB3* | rs145502455 | chr11:64263558 | A and G | 0.4 (2,028) | p.V806I (I) | 0.13 (0.021) | $1.6\times10^{-10}$ | 0.03 (0.020) | 0.11 |
| *FNIP1* | rs115209326 | chr5:131672891 | T and C | 0.3 (1,655) | p.R518Q (Q) | -0.13 (0.023) | $4.8\times10^{-9}$ | -0.12 (0.023) | $3.8\times10^{-7}$ |

Analyses are from 450,562 European ancestry individuals. Genomic coordinates according to human genome reference sequence hg38.
* Adjusting for conditionally-independent index variants highlighted in the joint conditional model (see Supplementary Table 3 for list of index variants at loci where fine-mapping supported causal role of these variants and Supplementary Table 4 for other loci).
‡ Variants in addition to the one of the sex-combined primary analysis which were identified in a secondary analysis in 244,478 women from the UK Biobank study ($p<5\times10^{-8}$).
Abbreviations: SE, standard error; WHR, waist-to-hip ratio; BMI, body mass index; SD, standard deviation.

**Supplementary Table 2. a. Correlation between the UK Biobank WES data and the genotype data for rare nonsynonymous variants with a minor allele frequency (MAF) between 0.1% - 0.5% in overlapping samples. b. Box plot of rare allele concordance between the UK Biobank WES data and the genotype data for rare nonsynonymous variants with a MAF between 0.1% - 0.5% in overlapping samples.**

**a.**

| Min | 1st Q | Median | Mean | 3rd Q | Max |
|---|---|---|---|---|---|
| 0.002 | 0.989 | 0.996 | 0.988 | 0.999 | 1 |

**b.**

| Min | 1st Q | Median | Mean | 3rd Q | Max |
|---|---|---|---|---|---|
| 0.002 | 0.991 | 0.997 | 0.987 | 0.999 | 1 |

**Supplementary Table 3: a. Conditionally independent index variants and fine-mapping at the *CALCRL*, *PLIN1*, *PDE3B* and *ACVR1C* loci.** Analyses are from 450,562 European ancestry individuals. Beta and standard errors are in standardized units of BMI-adjusted WHR per copy of the effect allele. Genomic coordinates according to human genome reference sequence GRCh38**. b. Formal conditional analyses at the *CALCRL*, *PLIN1*, *PDE3B* and *ACVR1C* loci.** Formal conditional analyses were conducted using individual-level genotype data in a subset of 350,721 unrelated European ancestry participants of UK Biobank. Because this is a subset of the discovery study, associations estimates differ from those presented in Supplementary Table 1.

**a.**

| **Locus** | **Signal** | **dbSNP rsID** | **Genomic coordinate, chromosome, position, effect allele, other allele (effect allele frequency, %)** | **Annotation** | **Beta (SE) from univariate analysis, in SD units** | **p-value univariate analysis** | **Beta (SE) from conditional analysis*, in SD units** | **p-value conditional analysis*** | **Genomic position of 99% credible set window, (width in number of base pairs)** | **Number of variants in the credible set** | **PPA for the index variant, %** |
|---|---|---|---|---|---|---|---|---|---|---|---|
| *PLIN1* | 1‡ | rs139271800 | chr15:89671546:G:A (0.1%) | *PLIN1* p.L90P | -0.21 (0.029) | $5.5\times10^{-13}$ | -0.21 (0.029) | $5.5\times10^{-13}$ | 89671546 (1) | 1 | >99% |
| *PDE3B* | 1‡ | rs150090666 | chr11:14843853:T:C (0.1%) | *PDE3B* p.R783X | -0.26 (0.032) | $1.4\times10^{-15}$ | -0.25 (0.032) | $6.2\times10^{-15}$ | 14843853 (1) | 1 | >99% |
| | 2 | rs2970332 | chr11:14338889:G: A (23.1%) | *RRAS2* intronic | -0.02 (0.002) | $9.9\times10^{-12}$ | -0.02 (0.002) | $6.3\times10^{-12}$ | 14236464-14667794 (431,331) | 20 | 23% |
| | 3 | rs79634051 | chr11:14540399:C: G (2.8%) | *PSMA1* intronic | -0.03 (0.006) | $6.4\times10^{-8}$ | -0.04 (0.006) | $2.1\times10^{-9}$ | 14221316-14869595 (648,280) | 15 | 78% |
| *ACVR1C* | 1 | rs55920843 | chr2:157556189:G: T (1.2%) | *ACVR1C* p.N150H | -0.08 (0.009) | $8.9\times10^{-19}$ | -0.09 (0.009) | $4.6\times10^{-20}$ | 157556189 (1) | 1 | >99% |
| | 2 | rs2444770 | chr2:157647227:C:T (14.8%) | 18kb 5' of *ACVR1C* | -0.02 (0.003) | $5.9\times10^{-13}$ | -0.02 (0.003) | $7.7\times10^{-15}$ | 157639990-157661726 (21,737) | 7 | 46% |
| | 3‡ | rs56188432 | chr2:157550353:G: A (0.2%) | *ACVR1C* p.I195T | -0.14 (0.021) | $4.9\times10^{-11}$ | -0.14 (0.021) | $5.4\times10^{-12}$ | 157550353 (1) | 1 | >99% |
| *CALCRL* | 1 | rs10177093 | chr2:187349092:G:T (45.6%) | *CALCRL* intronic | -0.02 (0.002) | $2.2\times10^{-27}$ | -0.02 (0.002) | $7.7\times10^{-26}$ | 187223800-187349092 (125,293) | 60 | 17% |
| | 2‡ | rs61739909 | chr2:187380712:G: A (0.3%) | *CALCRL* p.L87P | -0.13 (0.018) | $2.0\times10^{-13}$ | -0.12 (0.018) | $5.9\times10^{-12}$ | 187380712-187405532 (24,821) | 2 | 51% |

* Adjusting for conditionally-independent index variants highlighted in the joint conditional model.
‡ Variant identified in the genome-wide scan of rare nonsynonymous variants.

Abbreviations: SE, standard error; SD, standard deviation; PPA, posterior probability of association.

**b.**

| **Locus** | **Signal number** | **dbSNP rsID** | **Beta (SE) from univariate analysis** | **p-value univariate analysis** | **Beta (SE) from conditional analyses** | **p-value conditional analyses** |
|---|---|---|---|---|---|---|
| *CALCRL* | 1 | rs10177093 | -0.02 (0.002) | $4.4\times10^{-22}$ | -0.02 (0.002) | $2.2\times10^{-20}$ |
| | 2 | rs61739909 | -0.15 (0.021) | $9.8\times10^{-13}$ | -0.14 (0.021) | $5.5\times10^{-11}$ |
| *PLIN1* | 1 | rs139271800 | -0.16 (0.034) | $2.3\times10^{-6}$ | -0.16 (0.034) | $2.3\times10^{-6}$ |
| *PDE3B* | 1 | rs150090666 | -0.24 (0.037) | $2.0\times10^{-11}$ | -0.24 (0.037) | $1.7\times10^{-10}$ |
| | 2 | rs2970332 | -0.02 (0.003) | $9.8\times10^{-8}$ | -0.02 (0.003) | $1.2\times10^{-7}$ |
| | 3 | rs79634051 | -0.03 (0.007) | $2.9\times10^{-5}$ | -0.03 (0.007) | $2.0\times10^{-6}$ |
| *ACVR1C* | 1 | rs55920843 | -0.09 (0.011) | $3.1\times10^{-15}$ | -0.09 (0.011) | $3.0\times10^{-16}$ |
| | 2 | rs2444770 | -0.02 (0.003) | $2.8\times10^{-10}$ | -0.02 (0.003) | $1.7\times10^{-11}$ |
| | 3 | rs56188432 | -0.14 (0.025) | $9.9\times10^{-9}$ | -0.15 (0.025) | $4.7\times10^{-9}$ |

**Supplementary Table 4: Conditionally-independent index variants at the *ABHD15, PYGM, PLCB3 and FNIP1* regions.**

| Locus | dbSNP rsID | Chromosome and position |
|---|---|---|
| *Sex-combined* | | |
| *ABHD15* | rs62070804 | 17: 29562625 |
| *ABHD15* | rs561089333 | 17: 30422242 |
| *PYGM* | rs224170 | 11: 63897410 |
| *PYGM* | rs12419038 | 11: 64145264 |
| *PYGM* | rs3751122 | 11: 64185649 |
| *PYGM* | rs11231721 | 11: 64190363 |
| *PYGM* | rs7952318 | 11: 64193770 |
| *PYGM* | rs56271783 | 11: 64237251 |
| *PYGM* | rs75152214 | 11: 64543641 |
| *PYGM* | rs186402106 | 11: 64700853 |
| *PYGM* | rs2306363 | 11: 65638129 |
| *PYGM* | rs10750766 | 11: 65706327 |
| *PYGM* | rs4645917 | 11: 65714169 |
| *PYGM* | rs593982 | 11: 65745636 |
| *Sex-specific analysis in women* † | | |
| *PLCB3* | rs11231698 | 11: 64109691 |
| *PLCB3* | rs3751122 | 11: 64185649 |
| *PLCB3* | rs138055838 | 11: 64220169 |
| *PLCB3* | rs56271783 | 11: 64237251 |
| *PLCB3* | rs186826945 | 11: 64452647 |
| *FNIP1* | rs74667082 | 5: 131412611 |

Conditional analyses estimated the association of each index variant while adjusting for all other index variants at the region. Index variants were selected using a joint meta-analysis model with GCTA (1).

† Identified in a secondary analysis in 244,478 women from the UK Biobank study ($p<5\times10^{-8}$).

**Supplementary Table 5: STAAR-O (2) gene-based results for genes discovered in the single variant analysis.**

| Category | Gene | Number of Variants | Pval | Beta | SE |
|---|---|---|---|---|---|
| pLoF | *PLIN1* | 31 | $9.82\times10^{-9}$ | -0.27 | 0.05 |
| Moderate | *PLIN1* | 216 | $9.21\times10^{-6}$ | -0.02 | 0.01 |
| pLoF + Moderate | *PLIN1* | 292 | $4.94\times10^{-6}$ | -0.03 | 0.01 |
| pLoF | *ACVR1C* | 9 | $5.50\times10^{-2}$ | -0.48 | 0.24 |
| Moderate | *ACVR1C* | 130 | $4.57\times10^{-7}$ | -0.15 | 0.03 |
| pLoF + Moderate | *ACVR1C* | 139 | $1.68\times10^{-7}$ | -0.15 | 0.03 |
| pLoF | *PDE3B* | 65 | $1.41\times10^{-6}$ | -0.21 | 0.04 |
| Moderate | *PDE3B* | 392 | $8.12\times10^{-2}$ | -0.007 | 0.01 |
| pLoF + Moderate | *PDE3B* | 437 | $2.26\times10^{-5}$ | -0.03 | 0.01 |
| pLoF | *CALCRL* | 19 | $1.10\times10^{-1}$ | 0.31 | 0.14 |
| Moderate | *CALCRL* | 114 | $1.12\times10^{-3}$ | -0.07 | 0.02 |
| pLoF + Moderate | *CALCRL* | 133 | $1.16\times10^{-3}$ | -0.06 | 0.02 |

Abbreviations: pLOF, predicted loss of function; Pval, STAAR-O p-value; Beta, effect size; SE, standard error.

**Supplementary Table 6: Significant gene-based results from the exome-wide scan for $WHR_{adjBMI}$.**

| Category | Gene | Number of variants | Genotype counts (RR/RA/AA) | Pval | Beta | SE |
|---|---|---|---|---|---|---|
| pLoF | *PLIN4* | 65 | 183,900/1,065/0 | $5.86\times10^{-7}$ | 0.16 | 0.03 |
| pLoF | *PLIN1* | 31 | 184,572/388/5 | $9.82\times10^{-9}$ | -0.27 | 0.05 |
| pLoF | *INSR* | 27 | 184,904/61/0 | $6.21\times10^{-7}$ | -0.64 | 0.12 |
| Moderate | *ACVR1C* | 130 | 183,551/1,414/0 | $4.57\times10^{-7}$ | -0.15 | 0.03 |
| pLoF + Moderate | *ACVR1C* | 139 | 183,535/1,430/0 | $1.68\times10^{-7}$ | -0.15 | 0.03 |
| pLoF | *PDE3B* | 44 | 184,474/491/0 | $1.41\times10^{-6}$ | -0.21 | 0.04 |

Abbreviations: pLOF, predicted loss of function; MAF, minor allele frequency; Pval, STAAR-O p-value; Beta, effect size; SE, standard error; RR, reference/reference genotype; RA, reference/alternate genotypes; AA, alternate/alternate genotypes.

**Supplementary Table 7: Sex-specific results for gene-based analysis.**

| Gene | | | Women (n=101,569) | | | Men (n=82,677) | | |
|---|---|---|---|---|---|---|---|---|
| **Category** | **Gene** | **P_sexdiff** | **Beta** | **SE** | **Pval** | **Beta** | **SE** | **Pval** |
| pLoF | *PLIN4* | $2.08\times10^{-2}$ | 0.214 | 0.039 | $1.66\times10^{-7}$ | 0.073 | 0.046 | $1.55\times10^{-1}$ |
| pLoF | *PLIN1* | $2.83\times10^{-1}$ | -0.330 | 0.065 | $8.96\times10^{-8}$ | -0.223 | 0.075 | $4.56\times10^{-3}$ |
| pLoF | *INSR* | $4.62\times10^{-7}$ | -1.218 | 0.168 | $1.44\times10^{-12}$ | 0.046 | 0.186 | $7.53\times10^{-1}$ |
| Moderate | *ACVR1C* | $1.13\times10^{-1}$ | -0.181 | 0.035 | $7.78\times10^{-7}$ | -0.098 | 0.039 | $5.28\times10^{-2}$ |
| pLoF + Moderate | *ACVR1C* | $1.22\times10^{-1}$ | -0.183 | 0.034 | $4.60\times10^{-7}$ | -0.103 | 0.039 | $4.03\times10^{-2}$ |
| pLoF | *PDE3B* | $1.85\times10^{-3}$ | -0.334 | 0.059 | $5.04\times10^{-8}$ | -0.057 | 0.067 | $1.87\times10^{-1}$ |

Abbreviations: pLOF, predicted loss of function; P_sexdiff, p-value for the significance of the difference in women and men beta values; Pval, STAAR-O p-value; Beta, effect size; SE, standard error; RR, reference/reference genotype; RA, reference/alternate genotypes; AA alternate/alternate genotypes.

**Supplementary Table 8: Quality control measurements for variants included in the refined gene-based tests and single marker association results for all included variants.**

[Excel file: SupplementaryTable8_WHRadjBMI.xlsx]

**Supplementary Table 9: a. Summary statistics for quantitative traits from gene-based association analyses in sex-combined, women-only and male-only data. b. Summary statistics for binary traits from gene-based association analyses in sex-combined, women-only and male-only data.** Details for all phenotypes and are given on Supplementary Table 12.

[Excel file: SupplementaryTable9_WHRadjBMI.xlsx]

Abbreviations: pLOF, predicted loss of function; Pval, STAAR-O p-value for phenotypic traits in Supplementary Table 9a and generalized linear model for binary traits Supplementary Table 9b; Beta, effect size; SE, standard error; Beta_LCI, lower 95% confidence interval for effect size in; Beta_UCI, upper 95% confidence interval for effect size; TG, triglyceride; HDL, high-density lipoprotein; LDL, low-density lipoprotein.

**Supplementary Table 10: a. Lookup from Type 2 Diabetes Knowledge Portal for type 2 diabetes.** Seven different masks were applied to filter the variants used in the association analysis: LofTee (predicted loss of function); 5/5 (predicted deleterious by 5 methods); 16/16 (predicted deleterious by 16 methods); 5/5 + LofTee LC (predicted deleterious by 5 methods, plus LofTee low confidence), 5/5 + 0/5 1% (variants predicted deleterious by 5 methods, plus variants with minor allele frequency < 1% that are not predicted to be deleterious by any of 5 methods); 5/5 + 1/5 1% (variants predicted deleterious by 5 methods, plus variants with minor allele frequency < 1% that are predicted to be deleterious by 1 of 5 methods); and 11/11 (predicted deleterious by 11 methods). Rows in bold have P-value ≤ 0.05. Accessed from https://t2d.hugeamp.org/ on 02/09/2021. **b. Lookup from AstraZeneca PheWAS Portal (3) for type 2 diabetes, non-insulin-dependent diabetes mellitus (Union#E11#E11) and chronic ischaemic heart disease (Union#I25#I25).** The variant categories used in collapsing models are provided in the parentheses in 'Collapsing Model' column. Rows in bold have P-value ≤ 0.05. Accessed from https://azphewas.com/ on 02/09/2021.

**a.**

| Gene | Mask | P-value | Combined AF | Passing Variants | Singleton Variants | Standard Error | Sample Size | Odds Ratio |
|---|---|---|---|---|---|---|---|---|
| ***PLIN4*** | **LofTee** | **0.042** | **0.0042** | **17** | **6** | **0.153** | **43,125** | **1.3641** |
| ***PLIN4*** | **16/16** | **0.042** | **0.0042** | **17** | **6** | **0.153** | **43,125** | **1.3641** |
| ***PLIN4*** | **11/11** | **0.042** | **0.0042** | **17** | **6** | **0.153** | **43,125** | **1.3641** |
| ***PLIN4*** | 5/5 | 0.907 | 0.0115 | 24 | 8 | 0.094 | 43,125 | 1.0110 |
| ***PLIN4*** | 5/5 + LofTee LC | 0.907 | 0.0115 | 24 | 8 | 0.094 | 43,125 | 1.0110 |
| ***PLIN4*** | 5/5 + 1/5 1% | 0.793 | 0.0350 | 174 | 59 | 0.051 | 43,125 | 1.0134 |
| ***PLIN4*** | 5/5 + 0/5 1% | 0.653 | 0.0547 | 307 | 111 | 0.039 | 43,125 | 0.9825 |
| ***PLIN1*** | 11/11 | 0.478 | 0.0009 | 11 | 5 | 0.337 | 43,125 | 0.7905 |
| ***PLIN1*** | 5/5 | 0.237 | 0.0033 | 19 | 10 | 0.171 | 43,125 | 0.8178 |
| ***PLIN1*** | 5/5 + LofTee LC | 0.237 | 0.0033 | 19 | 10 | 0.171 | 43,125 | 0.8178 |
| ***PLIN1*** | 5/5 + 1/5 1% | 0.928 | 0.0121 | 87 | 39 | 0.089 | 43,125 | 0.9920 |
| ***PLIN1*** | 5/5 + 0/5 1% | 0.678 | 0.0153 | 132 | 56 | 0.079 | 43,125 | 0.9679 |
| ***INSR*** | **LofTee** | **0.017** | **0.0003** | **11** | **10** | **0.595** | **43,125** | **3.6645** |
| ***INSR*** | **16/16** | **0.015** | **0.0004** | **13** | **11** | **0.536** | **43,125** | **3.3304** |
| ***INSR*** | 11/11 | 0.229 | 0.0008 | 24 | 18 | 0.343 | 43,125 | 1.4992 |
| ***INSR*** | 5/5 | 0.836 | 0.0012 | 32 | 24 | 0.282 | 43,125 | 1.0596 |
| ***INSR*** | 5/5 + LofTee LC | 0.836 | 0.0012 | 32 | 24 | 0.282 | 43,125 | 1.0596 |
| ***INSR*** | 5/5 + 1/5 1% | 0.814 | 0.0234 | 212 | 128 | 0.064 | 43,125 | 0.9850 |

| | | | | | | | | |
|---|---|---|---|---|---|---|---|---|
| ***INSR*** | 5/5 + 0/5 1% | 0.803 | 0.0284 | 251 | 145 | 0.058 | 43,125 | 0.9855 |
| ***ACVR1C*** | LofTee | 0.534 | 0.0000 | 2 | 2 | 2.112 | 43,125 | 0.3845 |
| ***ACVR1C*** | 16/16 | 0.056 | 0.0001 | 5 | 5 | 1.654 | 43,125 | 0.1131 |
| ***ACVR1C*** | 11/11 | 0.084 | 0.0019 | 14 | 10 | 0.240 | 43,125 | 0.6662 |
| ***ACVR1C*** | 5/5 | 0.053 | 0.0020 | 15 | 10 | 0.238 | 43,125 | 0.6380 |
| ***ACVR1C*** | 5/5 + LofTee LC | 0.053 | 0.0020 | 15 | 10 | 0.238 | 43,125 | 0.6380 |
| ***ACVR1C*** | 5/5 + 1/5 1% | 0.152 | 0.0127 | 67 | 40 | 0.090 | 43,125 | 0.8791 |
| ***ACVR1C*** | 5/5 + 0/5 1% | 0.119 | 0.0132 | 79 | 47 | 0.089 | 43,125 | 0.8717 |
| ***PDE3B*** | LofTee | 0.562 | 0.0010 | 12 | 9 | 0.316 | 43,125 | 0.8349 |
| ***PDE3B*** | 16/16 | 0.678 | 0.0010 | 13 | 10 | 0.311 | 43,125 | 0.8802 |
| ***PDE3B*** | 11/11 | 0.739 | 0.0013 | 21 | 17 | 0.280 | 43,125 | 0.9118 |
| ***PDE3B*** | 5/5 | 0.847 | 0.0020 | 37 | 27 | 0.222 | 43,125 | 0.9584 |
| ***PDE3B*** | 5/5 + LofTee LC | 0.766 | 0.0020 | 38 | 28 | 0.221 | 43,125 | 0.9369 |
| ***PDE3B*** | 5/5 + 1/5 1% | 0.770 | 0.0238 | 178 | 98 | 0.064 | 43,125 | 0.9815 |
| ***PDE3B*** | 5/5 + 0/5 1% | 0.829 | 0.0331 | 263 | 148 | 0.054 | 43,125 | 1.0116 |

**b.**

| Gene | Phenotype | Collapsing model (Explanation) | P value | No. participants | No. cases with QV | No. controls with QV | Odds ratio | Odds ratio LCI | Odds ratio UCI |
|---|---|---|---|---|---|---|---|---|---|
| *PLIN4* | Type 2 diabetes | Synonymous negative control (synonymous variants with MAF≤0.05%) | $8.69\times10^{-2}$ | 162,620 | 14 | 844 | 1.60 | 0.94 | 2.72 |
| *PLIN1* | Type 2 diabetes | NA | NA | NA | NA | NA | NA | NA | NA |
| *INSR* | Type 2 diabetes | NA | NA | NA | NA | NA | NA | NA | NA |
| *ACVR1C* | Type 2 diabetes | Ultra-rare damaging (non-synonymous variants with MAF≤0.005%, REVEL score ≥ 0.25) | $6.95\times10^{-2}$ | 162,620 | 3 | 89 | 3.25 | 1.03 | 10.28 |
| *PDE3B* | Type 2 diabetes | Non-synonymous recessive (non-synonymous variants with MAF≤1%) | $9.23\times10^{-2}$ | 162,620 | 3 | 101 | 2.86 | 0.91 | 9.04 |
| *PLIN1* | Non-insulin-dependent diabetes mellitus (Union#E11#E11) | NA | NA | NA | NA | NA | NA | NA | NA |
| *PLIN4* | Non-insulin-dependent diabetes mellitus (Union#E11#E11) | NA | NA | NA | NA | NA | NA | NA | NA |
| *INSR* | Non-insulin-dependent diabetes mellitus (Union#E11#E11) | NA | NA | NA | NA | NA | NA | NA | NA |
| *ACVR1C* | Non-insulin-dependent diabetes mellitus (Union#E11#E11) | NA | NA | NA | NA | NA | NA | NA | NA |
| *PDE3B* | Non-insulin-dependent diabetes mellitus (Union#E11#E11) | Non-synonymous recessive (non-synonymous variants with MAF≤1%) | $9.66\times10^{-2}$ | 201,921 | 18 | 107 | 1.54 | 0.94 | 2.54 |
| *PLIN4* | Chronic ischaemic heart disease (Union#I25#I25) | NA | NA | NA | NA | NA | NA | NA | NA |
| ***PLIN1*** | **Chronic ischaemic heart disease (Union#I25#I25)** | Flexible MAF, damaging non-synonymous (non-synonymous variants with MAF≤0.1%, REVEL score ≥ 0.25) | $2.67\times10^{-2}$ | 176170 | 48 | 424 | 0.71 | 0.53 | 0.96 |
| | | Flexible MAF, all non-synonymous (non-synonymous variants with MAF≤0.1%) | $1.88\times10^{-2}$ | 176170 | 179 | 1355 | 0.83 | 0.71 | 0.97 |
| | | Flexible MAF, non-synonymous, Missense Tolerance Ratio (MTR) informed (non-synonymous variants with MAF≤0.1%, MTR <25$^{th}$ %ile or intergenic MTR < 50$^{th}$ %ile) | $9.70\times10^{-3}$ | 176,170 | 88 | 740 | 0.75 | 0.60 | 0.93 |
| | | **Protein truncating (protein truncating variants with MAF≤0.1%)** | $\mathbf{4.49\times10^{-4}}$ | **176,170** | **22** | **284** | **0.49** | **0.32** | **0.75** |

| | | | | | | | | | |
|---|---|---|---|---|---|---|---|---|---|
| | | **Protein truncating (protein truncating variants with MAF≤5%)** | **$4.49\times10^{-4}$** | **176,170** | **22** | **284** | **0.49** | **0.32** | **0.75** |
| | | Protein truncating or rare damaging models combined (protein truncating variants with MAF≤5% and missense variants with MAF≤0.025% and REVEL score ≥ 0.25) | $4.64\times10^{-4}$ | 176,170 | 32 | 369 | 0.55 | 0.38 | 0.78 |
| *PLIN1* | Chronic ischaemic heart disease (Union#I25#I25) | Non-synonymous recessive (non-synonymous variants with MAF≤1%) | $8.51\times10^{-2}$ | 176,170 | 12 | 129 | 0.59 | 0.32 | 1.06 |
| *INSR* | Chronic ischaemic heart disease (Union#I25#I25) | Rare damaging missense (missense variants with MAF≤0.025% and REVEL score ≥ 0.25) | $9.84\times10^{-2}$ | 176,170 | 133 | 715 | 1.17 | 0.97 | 1.41 |
| | | Rare damaging, MTR informed (missense variants with MAF≤0.025% and REVEL score ≥ 0.25, MTR <25th %ile or intergenic MTR < 50th %ile) | $5.06\times10^{-2}$ | 176,170 | 100 | 506 | 1.25 | 1.00 | 1.54 |
| *ACVR1C* | Chronic ischaemic heart disease (Union#I25#I25)) | NA | NA | NA | NA | NA | NA | NA | NA |
| *PDE3B* | Chronic ischaemic heart disease (Union#I25#I25) | Rare damaging, MTR informed (missense variants with MAF≤0.025% and REVEL score ≥ 0.25, MTR <25th %ile or intergenic MTR < 50th %ile) | $4.98\times10^{-2}$ | 176,170 | 38 | 335 | 0.71 | 0.51 | 1.00 |
| | | Non-synonymous recessive (non-synonymous variants with MAF≤1%) | $8.57\times10^{-2}$ | 176,170 | 8 | 96 | 0.52 | 0.25 | 1.08 |

Abbreviations: QV, qualifying variant; LCI, lower 95% confidence interval; UCI, upper 95% confidence interval; NA, not available.

**Supplementary Table 11: Leave one out analysis results for the most significant single variant in significant genes.**

| Category | Gene | Variant | chr | pos | Major Allele | Minor Allele | Consequence | MAF | Gene-based p-value | Gene-based p value after dropping the variant |
|---|---|---|---|---|---|---|---|---|---|---|
| pLoF | *PLIN4* p.Q372X | rs201581703 | 19 | 4512804 | G | A | Stop gained | 0.158% | $5.86{\times}10^{-7}$ | $1.79{\times}10^{-4}$ |
| pLoF | *PLIN1* p.T338DfsX51 | rs750619494 | 15 | 89667122 | CTTCTGC AGGGT | C | Frameshift variant | 0.029% | $9.82{\times}10^{-9}$ | $9.29{\times}10^{-4}$ |
| pLoF | *INSR* p.525RX | rs1599937180 | 19 | 7168005 | G | A | Stop gained | 0.001% | 6.21×10-$^{7}$ | $2.61{\times}10^{-4}$ |
| Moderate | *ACVR1C* p.I195T | rs56188432 | 2 | 15755035 3 | A | G | Missense variant | 0.208% | $4.57{\times}10^{-7}$ | 0.026 |
| pLoF+ Moderate | *ACVR1C* p.I195T | rs56188432 | 2 | 15755035 3 | A | G | Missense variant | 0.208% | $1.68{\times}10^{-7}$ | 0.011 |
| pLoF | *PDE3B* p.R783X | rs150090666 | 11 | 14843853 | C | T | Stop gained | 0.095% | $1.41{\times}10^{-6}$ | 0.493 |

Abbreviations: pLOF, predicted loss of function; chr, chromosome; pos, position (b38); MAF, minor allele frequency; Pval, STAAR-O p-value; Beta, effect size; SE, standard error; RR, reference/reference genotype; RA, reference/alternate genotypes; AA alternate/alternate genotypes.

**Supplementary Table 12. Phenotypes used for phenotypic associations in UK Biobank.** Sex (for sex-combined analyses), sequencing batch (50K vs. 150K), genotyping array, and 10 genetic principal components were included as covariates in association analyses.

| **Outcome** | **Cases, N** | **Non-cases (for case-control studies) or participants (for continuous traits studies) in sub-study, N** | **Adjustments for the phenotype** |
|---|---|---|---|
| BMI | NA | 184,291 | RB-INV transformed |
| Gynoid fat | NA | 178,143 | logn transformed, adjusted for age and logn total body fat, RB-INV of residuals within each sex separately |
| Android fat | NA | 178,143 | logn transformed, adjusted for age and logn total body fat, RB-INV of residuals within each sex separately |
| Leg fat | NA | 178,143 | logn transformed, adjusted for age and logn total body fat, RB-INV of residuals within each sex separately |
| Arm fat | NA | 178,143 | logn transformed, adjusted for age and logn total body fat, RB-INV of residuals within each sex separately |
| Trunk fat | NA | 178,143 | logn transformed, adjusted for age and logn total body fat, RB-INV of residuals within each sex separately |
| HbA1c | NA | 175,778 | RB-INV transform within aliquots, excluded prevalent T2D cases. |
| HDL cholesterol | NA | 161,239 | RB-INV transform within aliquots |
| LDL cholesterol | NA | 146,020 | RB-INV transform within aliquots, statin users were excluded |
| Triglycerides (TG) | NA | 175,271 | ln transformed, RB-INV transform within aliquots |
| TG/HDL ratio | NA | 161,102 | RB-INV transform within aliquots |
| Type 2 Diabetes | 12875 | 171,462 | According to the previously published UKBB probable T2D algorithm (27631769) based on baseline self-reported diabetes or medications, in addition to evidence from electronic health records (Hospital Episode Statistics or Death Registration) consistent with T2D (International Statistical Classification of Diseases and Related Health Problems Tenth Revision code E11) |
| Coronary Heart Disease | 11821 | 172,516 | Based on CALIBER working group's definition based on primary and secondary care records in UK Biobank |

Abbreviations: BMI, body mass index; HDL high-density lipoproteins; LDL low-density lipoproteins; RB-INV, rank-based inverse normal transformation; NA, not available.

**Supplementary Figure 1: Regional association plots of the overall and statistically-decomposed signals at the *CALCRL*, *PLIN1*, *PDE3B* and *ACVR1C* genes.** Plots were drawn using LocusZoom (4). Joint meta-analysis models using GCTA (1) were used at each locus to assess how many independent signals were present. Then, at each locus each signal was statistically-decomposed from others by estimating associations of all variants in the region adjusted for all other index variants at the region. Fine-mapping of each signal was performed using a Bayesian approach (5).

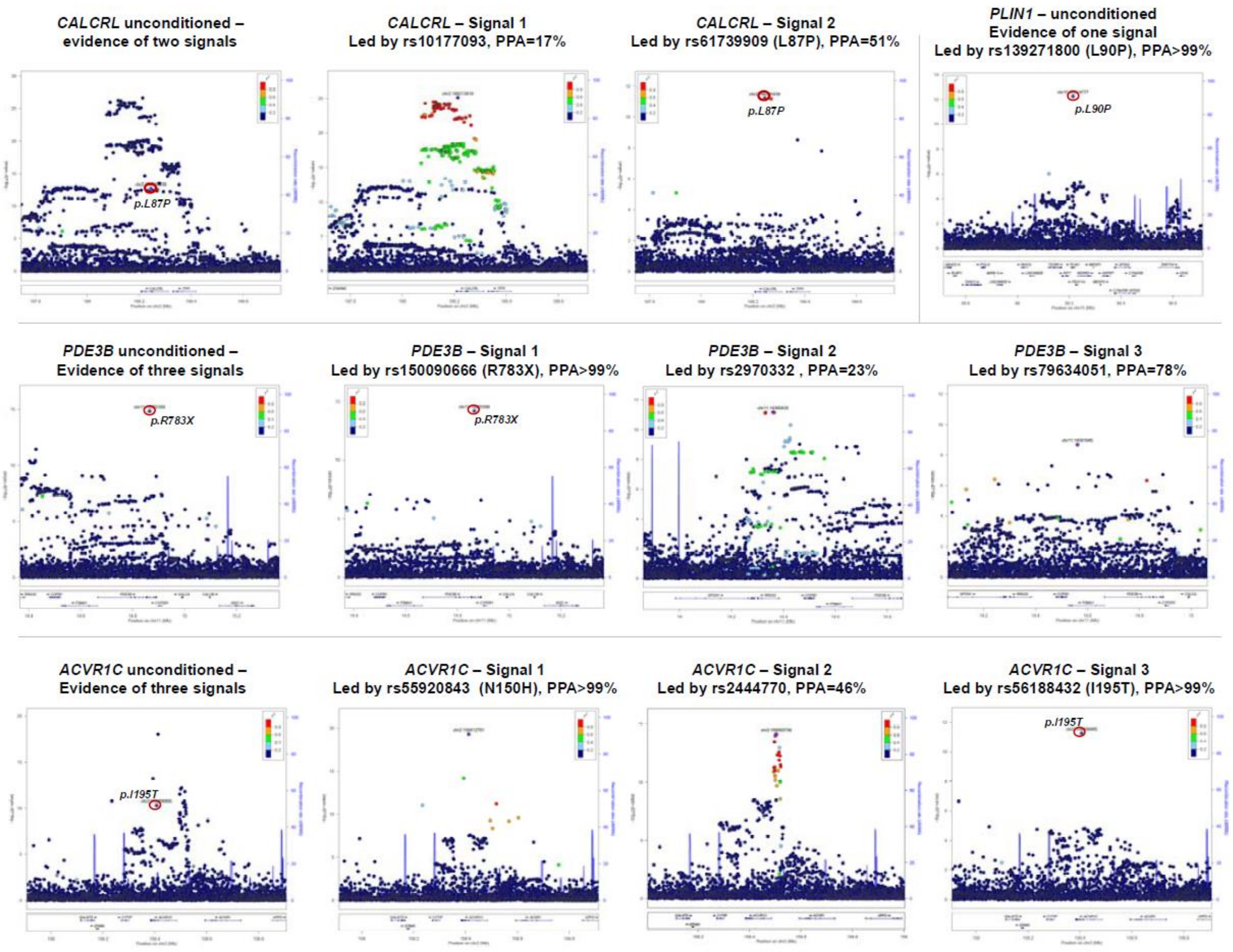

**Supplementary Figure 2: Association of the significant genes with other body fat distribution or cardiometabolic trait related phenotypes in UK Biobank for (a) continuous phenotypes and (b) binary phenotypes.** There was no rare predicted loss of function variant carriers for *INSR* in coronary heart disease (CHD) cases in women-only analyses. Abbreviations: pLoF, predicted loss of function; OR, odds ratio; WHRadjBMI; waist-to-hip ratio adjusted for body mass index; BMI, body mass index; TG, triglycerides; HD; high density lipoprotein, LDL low-density lipoprotein; TG/HDL, triglyceride to high density lipoprotein ratio; T2D, type 2 diabetes; CHD, coronary heart disease.

**a.**

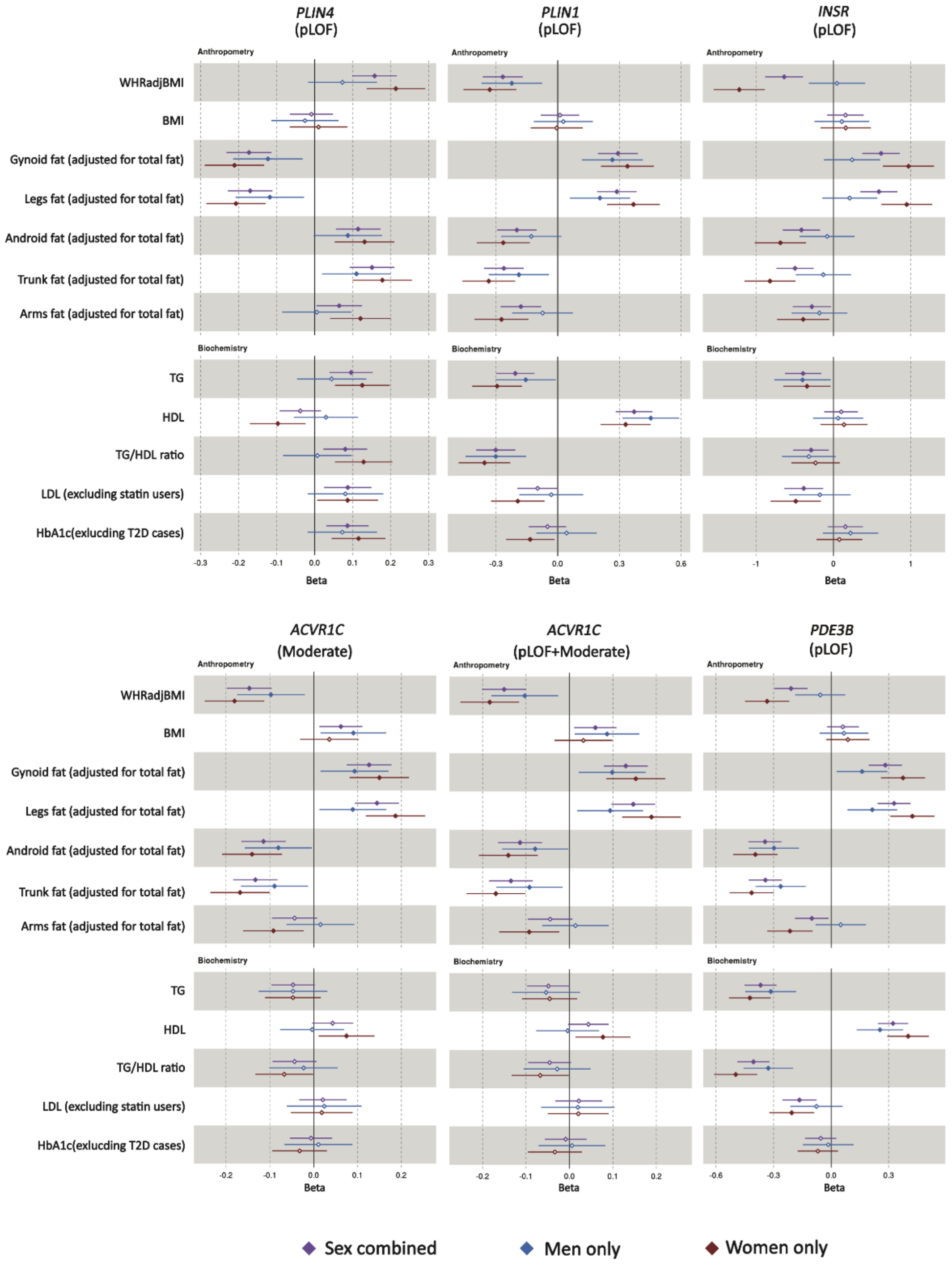

**b.**

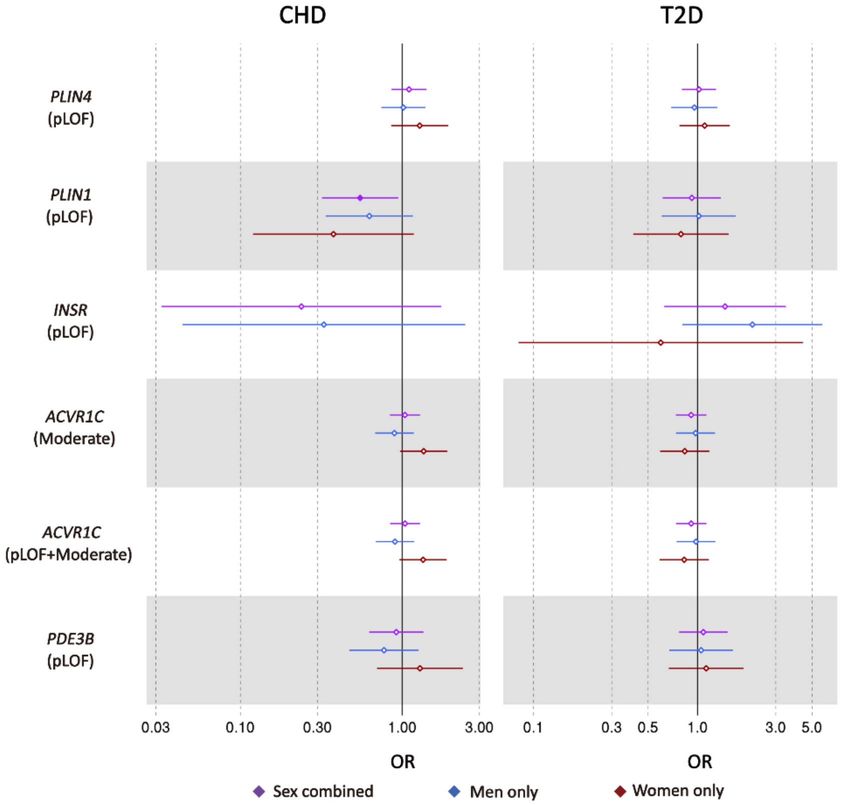
CHD
T2D
PLIN4
(pLOF)
PLIN1
(pLOF)
INSR
(pLOF)
ACVR1C
(Moderate)
ACVR1C
(pLOF+Moderate)
PDE3B
(pLOF)
0.03
0.10
0.30
1.00
3.00
0.1
0.3
0.5
1.0
3.0
5.0
OR
OR
Sex combined
Men only
Women only

**Supplementary Note 1: Genomic context analyses at the *PLIN1*, *ACVR1C*, *PDE3B* and *CALCRL* loci.**

Fine-mapping analyses provided strong statistical evidence for the causal association of rare nonsynonymous variants of *CALCRL*, *PLIN1*, *PDE3B* and *ACVR1C*.

At *PLIN1*, there was evidence of only one signal led by the rare p.L90P variant (rs139271800; Supplementary Figure 1), which was the only variant in the 99% credible set (PPA>99%; Supplementary Table 3).

At *ACVR1C*, there was evidence of three distinct signals (Supplementary Figure 1, Supplementary Table 3). The rare p.I195T variant led one of the secondary signals at this region and was the only variant in the 99% credible set (PPA>99%; Supplementary Table 3). In addition, the primary signal at this region was led by a low-frequency missense variant in *ACVR1C* (rs55920843, p.N150H), which also had the highest posterior probability in fine-mapping of this signal (PPA>99%; Table 1). Hence, fine-mapping of conditionally-independent signals at this locus converges on *ACVR1C* as causal gene for body fat distribution and p.I195T and p.N150H as causal variants for the respective association peaks.

At *PDE3B*, there was evidence of three signals, the strongest of which was led by the rs150090666 p.R783X nonsense variant in *PDE3B*, which was the only variant in the 99% credible set (PPA>99%; Supplementary Figure 1, Supplementary Table 3).

At *CALCRL*, there was evidence of two conditionally-independent signals (Supplementary Figure 1, Supplementary Table 3), led by the rs10177093 common variant and by the rare p.L87P variant, respectively. Fine-mapping at the latter signal yielded a 99% credible set including only two variants, rs61739909 (*CALCRL* p.L87P, posterior probability of casual association [PPA]=51%) and rs180960888 (intronic to *CALCRL*, PPA=48.5%). Hence, p.L87P is the most likely causal variant and *CALCRL* the most likely causal gene for this signal.

**Supplementary Note 2: Genomic context analyses at the *ABHD15*, *PYGM*, *PLCB3* and *FNIP1* loci.**

In the main analysis, we found associations of rare nonsynonymous variants in *ABHD15* and *PYGM* for which genomic context analyses were not consistent with a causal association. At *ABHD15*, conditional analyses revealed two distinct association signals tagged by the rs62070804 and rs561089333 index-variants respectively (Supplementary Table 4). The rare nonsynonymous rs141385558 p.G147D variant was not among these index variants and its association was greatly attenuated after adjusting for the two independent index variants ($p_{conditional}$=0.00019; Supplementary Table 1). At *PYGM*, there was a complex association pattern with evidence of up to 12 distinct signals (Supplementary Table 4). The rare nonsense rs116987552 p.R50X variant was not among these 12 index variants and its association was greatly attenuated after adjusting for these 12 index variants ($p_{conditional}$=0.00037; Supplementary Table 1). In the secondary analyses, we found associations of rare nonsynonymous variants in *PLCB3* (both in experiment-level p-value and sex-specific analyses) and *FNIP1* (sex-specific analyses) for which genomic context analyses were not consistent with a causal association. At *PLCB3*, there was evidence of up to ten independent signals (Supplementary Table 4) but the p.V806I variant was not among those and adjusting for the conditionally-independent index variants greatly attenuated its association ($p_{conditional}$>0.10; Supplementary Table 1). At *FNIP1* there was evidence of only one signal but the p.R518Q missense variant was not the lead variant (Supplementary Table 4). Adjusting for the lead variant attenuated the signal ($p_{conditional}=3.8\times10^{-7}$; Supplementary Table 1) and the variant had low posterior probability in the fine-mapping analysis (PPA=8.7%).

**Supplementary Note 3: Predictions regarding the variants identified through single marker analysis.**

<table>
<tr><th>Gene, allele<br>Gene product</th><th>Information on gene function, structural modelling and biological insights of associations reported in this study</th></tr>
<tr><td>*PLIN1* p.L90P<br>Perilipin 1</td><td>Perilipin 1 is a constitutive lipid droplet-associated protein predominantly expressed in adipocytes, where it is necessary for optimal triglyceride storage and for the precisely regulated release of fatty acids from the droplet (6). Perilipin 1 has a well-established role as a negative regulator of intracellular lipolysis (7, 8). Rare loss-of-function mutations in *PLIN1* cause autosomal dominant forms of partial lipodystrophy with lack of gluteo-femoral and leg fat, insulin resistance, dyslipidemia and type 2 diabetes (9). Leucine 90 is conserved in *Mammalia* and *Sauria* and replaced conservatively in lower species (*Inset Figure* 1). It lies at the N-terminal edge of the highly-conserved PAT domain responsible for the interaction with hormone sensitive lipase, the enzyme that catalyzes intracellular diglyceride hydrolysis (10). Although the structure of this region is unknown, it is predicted to be highly helical and the substitution of leucine with proline at position 90 is predicted to break the helix, introducing a sharp kink (*Inset Figure 2* ). Thus, p.L90P, which is associated with lower waist- to-hip ratio, higher overall adiposity, and lipid levels in our human genetic studies, may affect intracellular lipolysis by impacting on perilipin 1 interaction with hormone sensitive lipase. Our results show that nonsynonymous variation in this gene influences fat distribution and lipid levels in the general population, adding to the notion of shared genetic mechanisms between severe and subtle forms of human lipodystrophy (11).<br><br>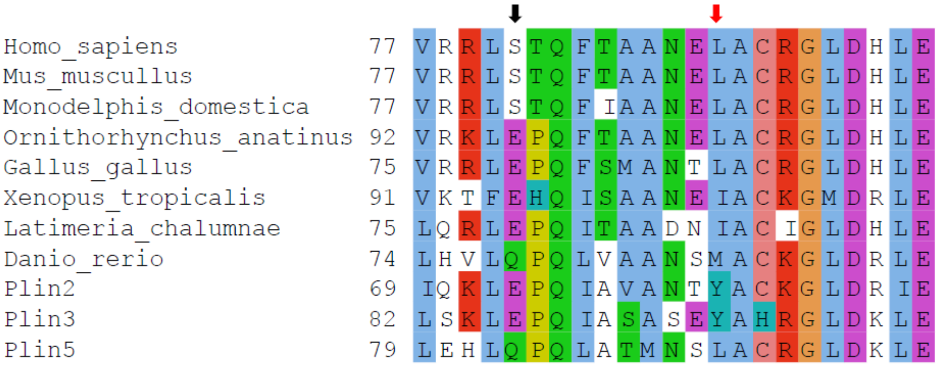<br><br>*Inset Figure 1*. Sequence alignment of perilipin 1 segments from representative species and human perilipin 2, 3 and 5. Perilipin 4 is not included due to divergence and multiplication of its N-terminus. The structure of the segment is modelled in *Inset Figure 2*. Arrows indicate the mutated L90 (red) and phosphorylated S81 (black).<br><br>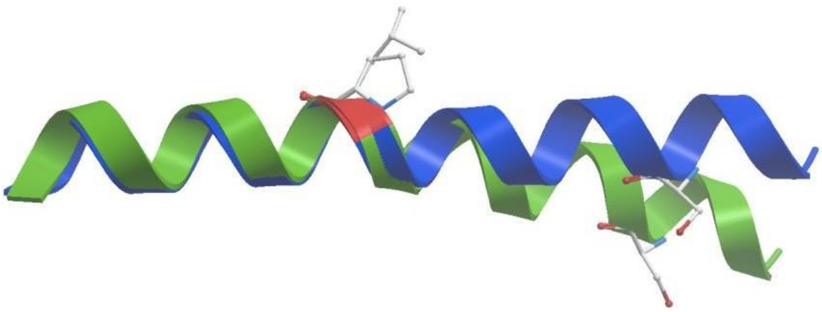<br>*Inset Figure 2*. Comparison of models of the native (blue) and mutated (green) fragment of perilipin 1 including amino acids 77 through 99. Built on the helical fragment of Protein Data Bank structure 4BJM/205-228 and superposed for the minimal root-mean-square deviation of the backbone (red) between L90 and P90. Their side chains are displayed together with S81 in ball and stick representation.</td></tr>
</table>

| | |
|---|---|
| *PDE3B* p.R783X<br>Phosphodiesterase 3B | Phosphodiesterase 3B is a membrane bound phosphodiesterase highly expressed in adipocytes, where it has been implicated in terminating intracellular lipolysis in response to insulin by degrading cyclic adenosine monophosphate (12). Phosphodiesterase 3B null mice manifest enhanced intracellular lipolysis, lower fat mass but higher insulin resistance (13). The premature stop codon generated by p.R783X falls within the proximal half of the catalytic domain of phosphodiesterase 3B, removing most of its $Mg^{2+}$ binding site (*Inset Figure 3*). Hence, this rare null variant, associated in our human genetic studies with lower waist-to-hip ratio, higher levels of peripheral adiposity and lower blood pressure and triglycerides is expected to result in a loss of catalytic function of phosphodiesterase 3B. If the protein is expressed, it could be embedded in the membrane through its intact N-terminal 6 membrane spanning domains and impair the phosphodiesterase 3B signalling complex (14) in a dominant- negative manner. Therefore, our data provide evidence of causal link between the loss of phosphodiesterase 3B function and greater peripheral fat, more favourable fat distribution and lower blood pressure and atherogenic lipids in humans. The effect size of this null allele on waist-to-hip ratio is over 6-fold greater than that of the strongest alleles found in GWAS of common variants (15).<br>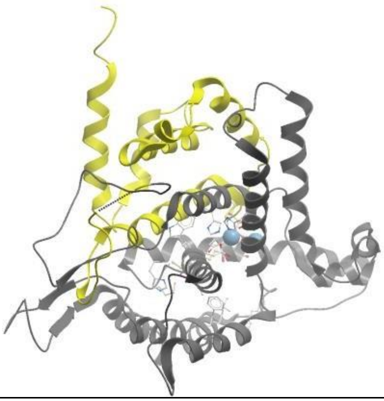<br>Inset Figure 3. Structure of the catalytic domain of PDE3B (Protein Data Bank coordinates 1SO2 (16)) with an inhibitor in the active site (Protein Data Bank coordinates 1ISO2((17))). The mutation R783X removes the protein part highlighted in grey. The N-terminus at the top of the figure is preceded by a 6 trans membrane helical domain. Inhibitor and active site residues are in ball and stick representation and the two blue balls indicate $Mg^{2+}$ ions. |
| *ACVR1C* p.I195T<br>Activin A Receptor Type 1C | The Activin A Receptor Type 1C is type I member of the family of transforming growth factor beta receptors transmitting signals from extracellular ligands to nuclear transcription. ACVR1C downregulates the key fat storage and glucose metabolism regulator peroxisome proliferator-activated receptor gamma (18). ACVR1C inhibits β-adrenergic signalling, mitochondrial biogenesis, lipid oxidation, and intracellular lipolysis in adipocytes (18). The *Inset Figure 4* shows a structural model of the intracellular part of ACVR1C. It can be seen that I195 forms a hinge between the N-terminal regulatory GS-domain and the kinase. Its side chain is tightly packed against both the kinase and the GS-domain. The serine/threonine epitope in the phosphorylation loop of the GS-domain is wedged in the active site between the kinase N- and C- lobes blocking access to the active site. Upon activation (phosphorylation), the GS-domain liberates the active site and interacts with its SMAD protein substrates. I195 is expected to be involved in any mutual GS- and kinase domain movement. Its side chain is buried in a hydrophobic environment and the change from aliphatic to polar residue should be significant. I195 is strictly conserved in all orthologues and in most metazoan paralogues including the 6 other human ones (*Inset Figure 5*). The rare replacements by different aliphatic amino acids (L, V) are very conservative. The mutation may influence kinase regulation as well as interaction with SMADs.<br>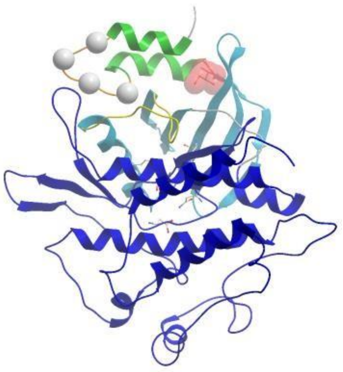<br>*Inset Figure 4*. Model of ACVR1C stabilized in its inactive conformation built according to the structure of TGFBR1 (Protein Data Bank coordinates 1B6C (19)). The N- terminus pointing to the membrane is on the top. The kinase domain is colored in blue, its N- and C-lobes distinguished in light and dark with the catalytic side- chains displayed in ball and stick and the activation loop in yellow. The regulatory GS-domain is in green with the phosphorylation epitope in orange and the alpha carbons of the residues to be phosphorylated upon activation highlighted by balls. The mutated L195 in ball and stick and space filling representation is in red. |

| | |
|---|---|
| | 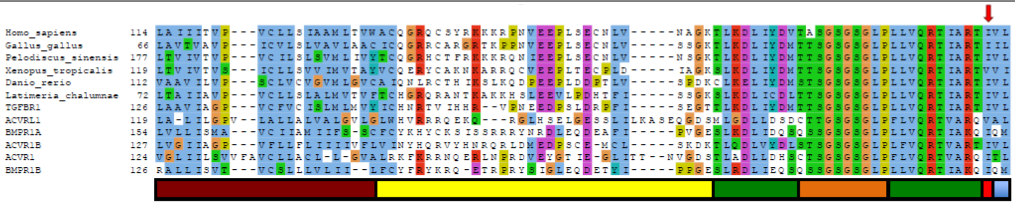<br>*Inset Figure 5.* Alignment of AVCR1C sequence segments from representative species and the human paralogues from TGF-beta family. The segment covers the transmembrane domain (brown), linker (yellow), GS-domain (green) with the phosphorylation loop (orange) followed by a hinge (red) and the first beta-strand of the kinase (blue). The latter four elements can be seen in 3D in *Inset Figure 4* using the same color coding. Arrow indicate the mutated I195. |
| *CALCRL* p.L87P<br>Calcitonin receptor-like | Calcitonin receptor-like receptor is a G-protein coupled receptor which requires association with one of the receptor activity-modifying proteins 1-3 (RAMP1-3) for ligand binding and receptor activation (20). When associated with RAMP1, it serves as a receptor for calcitonin gene-related peptide (CGRP), whereas when associated with RAMP2-3 it functions as a receptor for adrenomedullin which is most widely recognized as a vasoactive peptide (21, 22). Mouse knockouts of adrenomedullin (23) and CALCRL (24) are embryonic lethal. However, adrenomedullin, CALCRL and RAMP2-3 are all expressed in human adipocytes where adrenomedullin has been shown to stimulate intracellular lipolysis (25). Leucine 87 is not a conserved residue and is replaced by proline in several species (*Inset Figure 6*). The mutation is located at the tip of a strand in a beta-hairpin next to C88 and C127, both of which are strictly conserved (*Inset Figures 6-7*). The existence of the neighboring disulfide cross-link which is strictly conserved indicates that the orientation of the hairpin is important and it is indeed in close contact with RAMP2. The mutation may partially destabilize the beta-sheet, leading to a slight readjustment of the hairpin, and its interaction with adrenomedullin. The effect on ligand binding will however be indirect and probably mild in the complex with RAMP2 and adrenomedullin. However, the effect might be stronger when the receptor forms a complex with RAMP1 or 3 and interacts with different ligands. The association of p.L87P with lower waist-to-hip ratio coupled with the high expression of the *CALCRL* gene in adipose tissue point to a possible role in adipocyte biology of this G-protein coupled receptor.<br>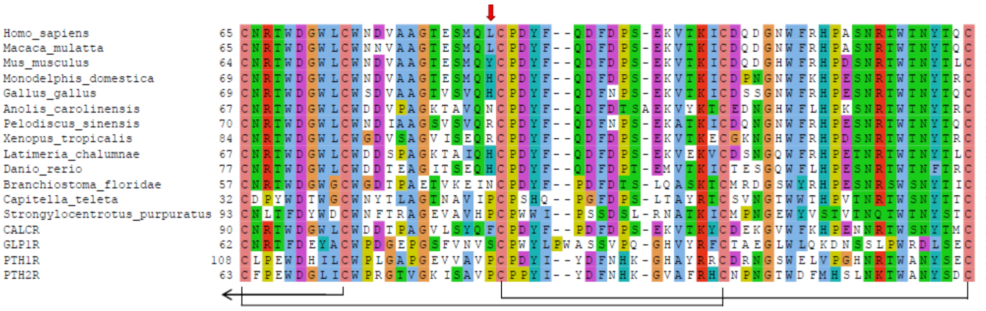<br>*Inset Figure 6.* Sequence alignment of CALCRL segments from representative species and the closest human homologues. The disulfide bonds are indicated by black connectors and the mutated L87 by a red arrow. The fragment covers the beta-barrel in *Inset Figure 7*. |

<table>
<tr>
<td></td>
<td>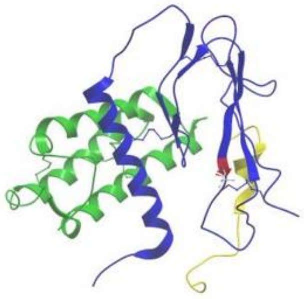

*Inset Figure 7*. Structure of the extracellular portion of the complex between CALCRL (blue), RAMP2 (yellow) and adrenomedullin (green). Extensions of the C-terminus of CALRL and the N-terminus of RAMP2 (not shown in this model) constitute the trans-membrane and intracellular domains of CALCRL and RAMP2. The position of L87 is in red and its side-chain in ball and stick representation. The side chains of the conserved structure stabilizing cysteines are also shown. Protein Data Bank coordinates 4RWF (26) were used for modelling.</td>
</tr>
</table>
